# Apathy precedes worsening depression, but not the reverse

**DOI:** 10.64898/2026.05.05.26352457

**Authors:** Sijia Zhao, Nadine Paans, Didi Rhebergen, Richard C. Oude Voshaar, Erik J. Giltay, Brenda W.J.H. Penninx, Masud Husain

## Abstract

Apathy–loss of motivation associated with diminished goal-directed behaviour–is among the most prevalent yet undertreated neuropsychiatric syndromes. Whether it represents a downstream residual manifestation of chronic depression or an independent and potentially upstream process remains unresolved. Here, leveraging two large-scale longitudinal psychiatric cohorts (N=2,482; age 24–93), we demonstrate that multidimensional apathy is structurally and conditionally independent from anhedonia, depressed mood, anxiety and fatigue, using item-level factor analysis and partial correlation networks. Crucially, in the 6-year longitudinal subsample (N=292), cross-lagged panel models revealed an asymmetric temporal precedence: apathy strongly predicted subsequent worsening of depressed mood and anhedonia, but not the reverse. This progressive motivational decline was not explained by underlying genetic neurodegenerative risk (APOE ε4 genotype) or cognitive decline. These findings overturn the assumption that motivational withdrawal follows loss of pleasure, and positions apathy as an upstream, independently targetable process in the evolution of depression.

## Introduction

Apathy is a pervasive neuropsychiatric syndrome that is highly prevalent across a wide range of brain disorders.^1,2^ In neurological conditions, such as Alzheimer’s disease, Parkinson’s disease and frontotemporal dementia, apathy is well-established as being associated with increased functional impairment and caregiver burden.^3,4^ Apathy is also common in psychiatric disorders. In major depression, 40–60% of patients meet established criteria for clinical apathy.^5,6^ However, the relationship between depression and apathy remains to be fully established. One way to examine this is by considering components of motivated behaviour.

Motivational deficits in depressed individuals have sometimes been subsumed under the symptom of anhedonia (loss of consummatory pleasure) or viewed as a residual manifestation of chronic depressive disorder.^7,8^ Preclinical models and human neuroimaging consistently demonstrate that the neural circuits governing ‘wanting’ (motivation to allocate effort, heavily dependent on dopaminergic projections to the striatum and anterior cingulate cortex) are biologically dissociable from those governing ‘liking’ (hedonic consummatory pleasure).^9,10^ While these circuit-level distinctions are increasingly recognised, applying them to human psychiatric taxonomies remains challenging.^1^ Consequently, apathy and depression are frequently treated as overlapping constructs in both clinical practice and intervention trials.^11^ However, high clinical comorbidity does not equate to construct identity.^6,12,13^ Indeed, conflating apathy with depression may help to explain why a substantial subset of depressed patients with prominent motivational deficits do not fully respond to standard serotonergic antidepressants, highlighting the urgent clinical need to better disentangle these symptom dimensions.^14,15^

A potentially informative way to clarify the relationship between constructs such as apathy, anhedonia and depression would be to test their temporal relationship in primary psychiatric populations. To the best of our knowledge such an investigation has never previously been performed. One possibility is that effortful, goal-directed behaviour is diminished in patients primarily because they have lost the capacity to experience pleasure.^16,17^ If this were the case, motivational withdrawal would be a downstream consequence of anhedonia or depression. Recent advances in translational neuroscience, however, invite a re-evaluation.

If the distributed frontostriatal circuits governing effort and motivation are particularly vulnerable to systemic and allostatic stress, motivational decline might actually emerge *before* profound mood disturbances.^10,18^ In this scenario, isolated apathy would not necessarily be a late-stage residual symptom of past depression, but could instead serve as an upstream, transdiagnostic indicator of impending affective decline. Establishing whether a reduction in the drive to act precedes the emergence of anhedonia or depressed mood would provide crucial insights into syndromic trajectories, potentially identifying apathy as an earlier, distinct target for intervention.

To empirically test this, we investigated the structural validity and longitudinal dynamics of apathy, anhedonia, depression, and anxiety across the adult lifespan. We leveraged two complementary longitudinal psychiatric cohorts: the Netherlands Study of Depression and Anxiety (NESDA), which provides a large age-diverse sample, and the Netherlands Study of Depression in Older Persons (NESDO), which provides repeated apathy assessments across baseline, 2-year and 6-year follow-up.^19–21^ The entire sample enabled high-powered item-level factor analysis, partial correlation networks and clinical profiling to test whether apathy is structurally separable from anhedonia, depressed mood and anxious arousal. The three-wave older-adult cohort then enabled cross-lagged panel modelling to test temporal precedence over six years. This design deliberately separated two inferential questions: whether apathy is an independent construct, and whether it precedes rather than follows worsening affective symptoms. We hypothesised that apathy forms a structurally distinct clinical phenotype that precedes, rather than follows, the progressive worsening of depressive symptoms over time.

## Results

### Apathy is highly prevalent in depression and ageing, with additive but non-synergistic effects

To investigate the structural and temporal dynamics of apathy across the adult lifespan, we leveraged data from 2,482 adults with depressive or anxiety disorders pooled from two independent cohorts (NESDA n = 2,011; NESDO n = 471). Baseline demographic and clinical characteristics for both cohorts are detailed in **Table 1** (see Methods). Within this pooled sample, the prevalence of clinical apathy (SAS score ≥ 14) increased alongside both advancing age and the ongoing presence of depressive symptoms (**Figure 1A**). Among participants without current depression, clinical apathy remained below 15% until age 60, before rising to approximately 45% by age 80. Conversely, among those experiencing moderate-to-severe depression, apathy prevalence exceeded 30% across all age groups and approached 95% by age 80.

**Figure 1:**
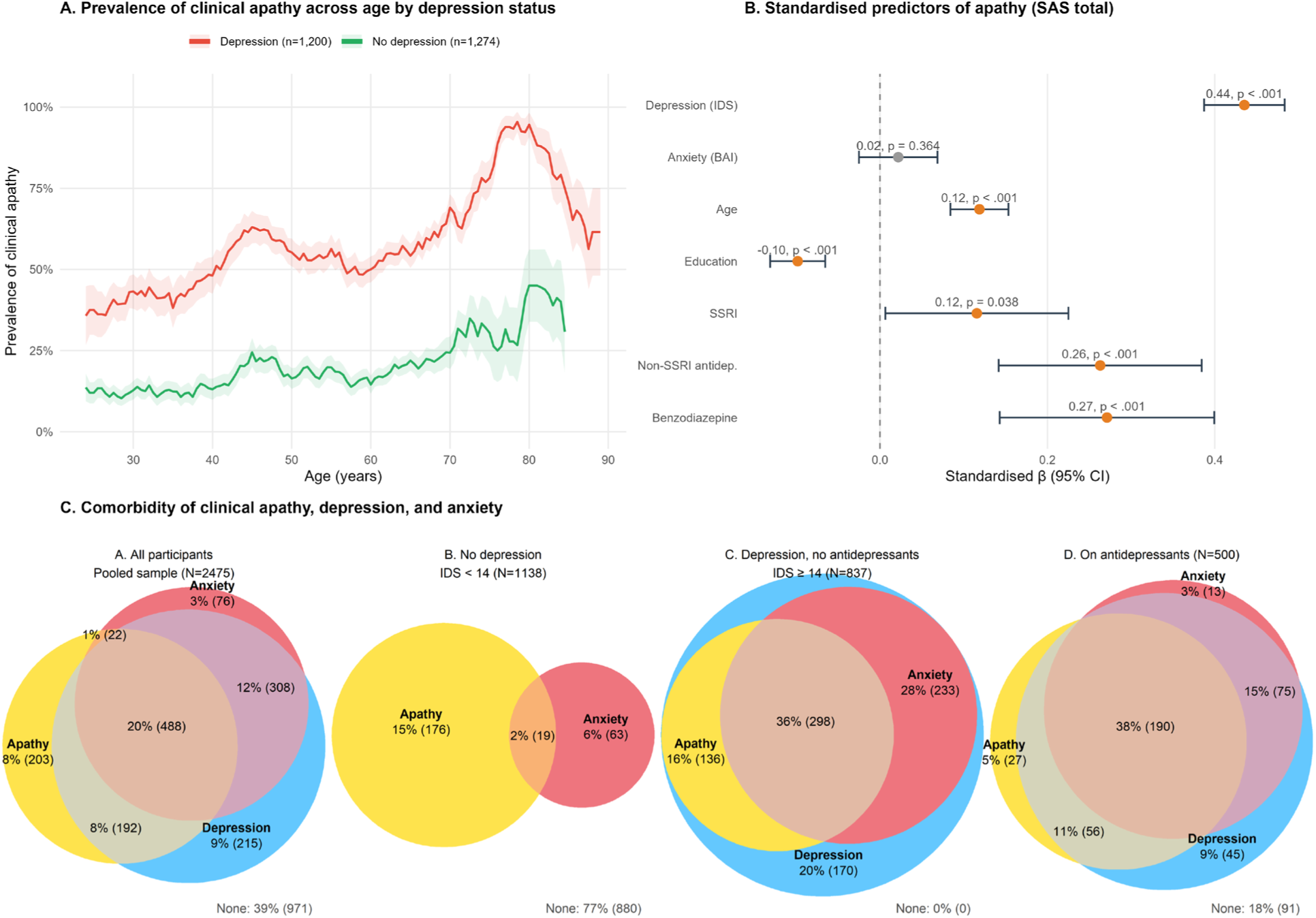
Clinical apathy by depression status across the adult lifespan. Pooled NESDA and NESDO sample (N = 2,482; age 24–94). (A) Prevalence of clinical apathy (SAS ≥ 14) across age, stratified by depression status (IDS-SR severity ≥ 14), using a 5-year sliding window (±2.5 yr; band = ±1 SEM). (See Supplementary Figure S1A for running mean SAS total score across age.) (B) Standardised regression coefficients from linear regression predicting continuous SAS (see Supplementary Figure S1B for odds ratios from logistic regression predicting clinical apathy, and Figures S1C–D for analysis in NESDO only with MMSE). (C) Comorbidity of clinical apathy, depression, and anxiety. Venn diagram showing the overlap of clinical apathy (SAS ≥ 14), depression (IDS-SR ≥ 14), and clinical anxiety (BAI ≥ 10) in the pooled sample (N = 2,475). (i) All participants. (ii) No depression, not on antidepressants (n = 1,138). (iii) Depression, not on antidepressants (n = 837). (iv) On antidepressants (n = 500).

**Table 1:** Demographic, clinical, and health characteristics of the NESDO and NESDA samples. NESDO is shown across all three waves because the SAS was administered at every assessment, enabling the longitudinal analyses; NESDA is shown only at Wave E because the SAS was administered only at this wave in NESDA. SAS = Starkstein Apathy Scale (clinical apathy ≥ 14). BAI = Beck Anxiety Inventory; severity categories (Kabacoff): normal (0–9), mild (10–18), moderate (19–29), severe (≥30). IDS-SR = Inventory of Depressive Symptomatology, Self-Report; severity categories: none (0–13), mild (14–25), moderate (26–38), severe (39–48), very severe (≥49). MMSE = Mini-Mental State Examination. WHODAS = WHO Disability Assessment Schedule 2.0. Dashes indicate measures not available at that wave.

| Characteristic | NESDO Baseline | NESDO 2-Year | NESDO 6-Year | NESDA Wave E |
| --- | --- | --- | --- | --- |
| N | 510 | 299 | 401 | 2,256 |
| Retention from baseline, % | 100 | 58.6 | 78.6 | 75.7 |
| Demographics |  |  |  |  |
| Age, years, mean (SD) | 70.1 (7.5) | — | — | 47.8 (13.1) |
| Female sex, n (%) | 331 (64.9%) | — | — | 1,496 (66.3%) |
| Education level, mean (SD) | 6.2 (2.8) | — | — | 5.6 (2.0) |
| Clinical measures |  |  |  |  |
| Apathy (SAS total), mean (SD) | 15.4 (6.2) | 13.7 (6.8) | 14.9 (7.2) | 10.9 (5.1) |
| Clinical apathy (SAS $\geq 14$), n (%) | 295 (61.8%) | 145 (50.0%) | 221 (55.7%) | 616 (30.6%) |
| BAI total, mean (SD) | 14.0 (11.8) | 8.7 (9.1) | 10.9 (10.5) | 8.4 (8.5) |
| IDS-SR total, mean (SD) | 24.4 (15.2) | 16.3 (12.4) | 19.1 (13.5) | 15.2 (11.9) |
| MMSE, mean (SD) | 27.9 (1.9) | — | — | — |
| MMSE $< 24$, n (%) | 14 (2.8%) | — | — | — |
| WHODAS (disability), mean (SD) | 20.7 (14.0) | — | — | — |
| IDS-SR severity, n (%) |  |  |  |  |
| None | 151 (30.1%) | 144 (49.7%) | 172 (43.1%) | 1,154 (53.8%) |
| Mild | 114 (22.7%) | 89 (30.7%) | 112 (28.1%) | 585 (27.3%) |
| Moderate | 134 (26.7%) | 38 (13.1%) | 69 (17.3%) | 304 (14.2%) |
| Severe | 63 (12.5%) | 11 (3.8%) | 33 (8.3%) | 73 (3.4%) |
| Very severe | 40 (8.0%) | 8 (2.8%) | 13 (3.3%) | 30 (1.4%) |
| BAI severity, n (%) |  |  |  |  |
| Normal | 207 (43.5%) | 189 (65.4%) | 232 (58.3%) | 1,425 (66.4%) |
| Mild | 122 (25.6%) | 61 (21.1%) | 89 (22.4%) | 450 (21.0%) |
| Moderate | 86 (18.1%) | 29 (10.0%) | 50 (12.6%) | 196 (9.1%) |
| Severe | 61 (12.8%) | 10 (3.5%) | 27 (6.8%) | 74 (3.4%) |
| Medication use |  |  |  |  |
| Any antidepressant, n (%) | 280 (54.9%) | 83 (28.2%) | 106 (26.4%) | 456 (20.2%) |
| SSRI, n (%) | 107 (21.0%) | 35 (11.9%) | 54 (13.5%) | 268 (11.9%) |
| Other antidepressant, n (%) | 107 (21.0%) | 52 (17.7%) | 53 (13.2%) | 114 (5.1%) |
| Benzodiazepine, n (%) | 186 (36.5%) | 56 (19.0%) | 113 (28.2%) | 90 (4.0%) |
| Health |  |  |  |  |
| BMI (kg/m<sup>2</sup>), mean (SD) | 26.5 (4.3) | — | — | 26.2 (5.1) |
| Fasting glucose (mmol/L), mean (SD) | 5.9 (1.4) | — | — | 5.5 (1.1) |
| ApoE $\epsilon 4$ carrier, n (%) | 147 (30.1%) | — | — | 589 (31.3%) |
| Chronic diseases,<br>mean (SD) | 2.0 (1.4) | — | — | 0.8 (1.0) |

A multiple linear regression predicting continuous apathy severity (adjusted R^2^ = 0.327; **Figure 1B**) confirmed that depression severity was the strongest predictor (standardised β = 0.435, p < 0.001), while anxiety severity was not significant (std. β = 0.022, p = 0.364). Age showed a modest positive association (std. β = 0.119, p < 0.001), and higher education was protective (std. β = −0.098, p < 0.001). Notably, after controlling for depression and anxiety severity, the use of psychotropic medications—specifically non-SSRI antidepressants (std. β = 0.263) and benzodiazepines (std. β = 0.271)—remained strong predictors of apathy. Logistic regression corroborated these findings.

However, despite this substantial clinical overlap between apathy and depression (**Figure 1C**), the interaction between depression and age was not significant (β = 0.006, p = 0.343; **Table 2**), indicating that depression and ageing exert independent, additive effects on apathy rather than synergistic ones. Consequently, the apathy prevalence gap between depressed and non-depressed groups remains constant across the adult lifespan (**Figure 1A**). Furthermore, a sensitivity analysis in the older NESDO subsample confirmed that global cognition (Mini-Mental State Examination, MMSE) did not significantly predict apathy severity (b = -0.08, p = 0.530, Supplementary Figure S1C) or the likelihood of clinical apathy (OR = 1.01, p = 0.889; Supplementary Figure S1D), demonstrating that these motivational deficits cannot be attributed merely to general neurocognitive impairment.

**Table 2:** Statistical tests for the depression–apathy association. Chi-square test and odds ratios for the association between depression (IDS-SR total ≥ 14, i.e., mild severity or above) and clinical apathy (SAS ≥ 14) in the pooled sample (N = 2,482). The interaction model is a logistic regression predicting clinical apathy with depression status, age, and the depression × age product term as predictors. A non-significant interaction (p = 0.343) indicates that the effect of depression on apathy does not vary with age.

| Test | Statistic | p |
| --- | --- | --- |
| Chi-square | $\chi^2 = 398.98$ | < 0.001 |
| Unadjusted OR | OR = 6.04 [5.02, 7.25] | < 0.001 |
| Age-adjusted OR | OR = 5.54 [4.60, 6.68] | < 0.001 |
| Interaction: Depression | b = 1.387 | < 0.001 |
| Interaction: Age | b = 0.020 | < 0.001 |
| Interaction: Depression × Age | b = 0.006 | 0.343 |

### The distinct factor structure of apathy, depression, and anxiety is robust across cohorts and longitudinally stable

Clinical apathy demonstrated substantial overlap with depression and anxiety, with 32.4% (488/1,504) of symptomatic participants meeting criteria for all three syndromes (**Figure 1C**). Notably, the comorbidity of apathy and anxiety without depression was exceedingly rare (1.5%, **Figure 1C**). This contrasts sharply with the prevalence of comorbid apathy and depression without anxiety (192 of 1,504; 12.8%) and comorbid anxiety and depression without apathy (308 of 1,504; 20.5%; pairwise comorbidity rates: χ² = 237.84, df = 2, p < 0.001).

Because high comorbidity does not equate to construct identity – just as anxiety and depression remain distinct despite frequent co-occurrence – we performed an item-level exploratory factor analysis (EFA) in the older NESDO sample (N = 510) and a confirmatory factor analysis (CFA) in the age-diverse NESDA sample.

In the NESDO baseline sample (N = 510, age range = 60–93 years), sampling adequacy for the EFA was excellent (KMO = 0.944; Bartlett’s test Chi-square = 16,817, p < 0.001). Parallel analysis of the 62 items (1,000 simulated datasets; maximum likelihood extraction) identified 10 factors (scree plot in **Supplementary Figure S2**). Maximum likelihood extraction with oblimin rotation yielded a clean simple structure (**Figure 2**): only 4 of the 62 items cross-loaded (loadings >= 0.30 on more than one factor), and communalities ranged from 0.24 to 0.87 (median = 0.54).

**Figure 2:**
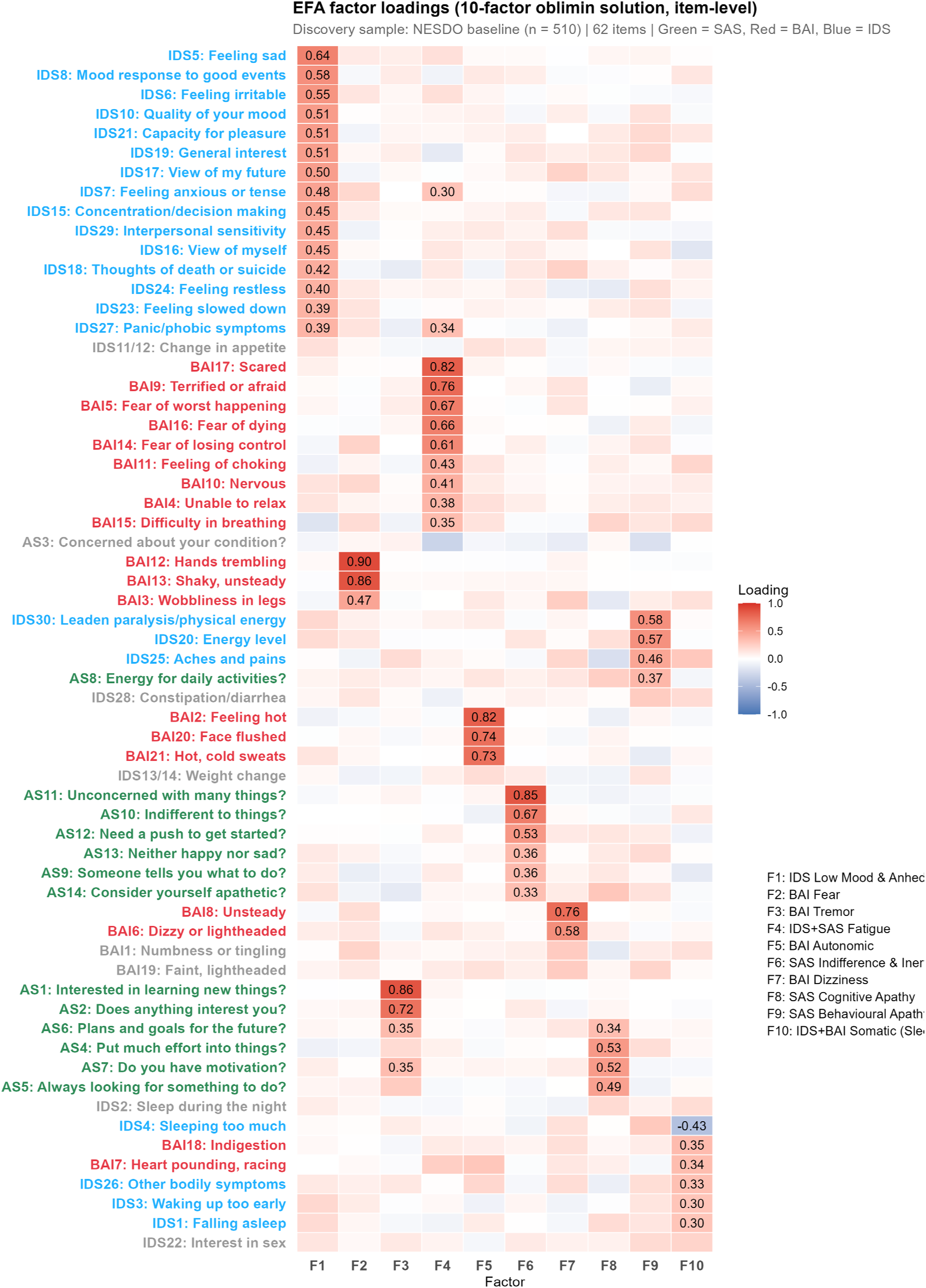
Exploratory factor analysis: 10-factor loading structure. Heatmap of factor loadings (maximum likelihood extraction, oblimin rotation) on 62 symptom items in the NESDO baseline sample (N = 510, mean age = 70.1 years, all with psychiatric conditions such as depression or anxiety). Three apathy factors (F6: Indifference & inertia, F8: Cognitive apathy, F9: Behavioural apathy) form a distinct cluster. F4 (IDS+SAS Fatigue) is the only cross-scale factor, loading both SAS and IDS items. Green = SAS, Red = BAI, Blue = IDS.

Three apathy factors (F6: Indifference and inertia; F8: Cognitive apathy; F9: Behavioural apathy) emerged as structurally distinct from depression (F1: Depressed mood and anhedonia) and anxiety (F2: Fear; F3: Tremor; F5: Autonomic arousal; F7: Dizziness). Crucially, F4 (Fatigue) loaded items from both the SAS and IDS, demonstrating that this structural separation is driven by true symptom content, not merely questionnaire variance. Full item-level loadings, communalities, and the factor correlation matrix are provided in **Supplementary Table S1 and Figure S3**.

This 10-factor model was confirmed via CFA in the independent NESDA sample (N = 2,256; RMSEA = 0.050, SRMR = 0.049), demonstrating that the structure is not sample-specific. Furthermore, longitudinal measurement invariance testing across the three NESDO waves confirmed configural, metric, and scalar invariance, ensuring that longitudinal variations in factor scores reflect true clinical change rather than measurement artefacts (**Table 3**).

**Table 3:** Longitudinal measurement invariance of the 10-factor model across three NESDO assessment waves. Fit indices for configural, metric (equal loadings), and scalar (equal intercepts) invariance models tested on the 10-factor structure (26 indicator items; F1–F10) across NESDO baseline, 2-year, and 6-year follow-up waves. ΔCFI and ΔRMSEA indicate change from the preceding model; invariance is supported when ΔCFI > −0.01. All three levels of invariance were supported, confirming that factor loadings and intercepts remained stable over the 6-year follow-up.

| Model | $\chi^2$ | df | CFI | TLI | RMSEA | SRMR | ΔCFI | ΔRMSEA | $\Delta\chi^2$ | Δdf | p | Supported |
| --- | --- | --- | --- | --- | --- | --- | --- | --- | --- | --- | --- | --- |
| Configural | 3580.9 | 1647 | .905 | .891 | .054 | .058 | — | — | — | — | — | Baseline |
| Metric | 3686.1 | 1699 | .902 | .891 | .054 | .061 | –.003 | < .001 | 105.2 | 52 | < .001 | Yes |
| Scalar | 3794.2 | 1751 | .900 | .892 | .054 | .062 | –.003 | < .001 | 108.1 | 52 | < .001 | Yes |

### Apathy does not correlate with anxiety, after controlling depression severity

To map the conditional dependencies between these constructs, we examined partial Spearman correlations. Zero-order correlations showed apathy was positively associated with both depression (ρ = 0.58, p<0.001) and anxiety (ρ = 0.45, p<0.001) across all subdomains. This pattern extended across all five apathy subscales of SAS defined by Pedersen et al. (Indifference, Cognitive, Behavioural, Motivation, Insight),^22^ with correlations ranging from weak to strong (**Figure 3A**). However, controlling for depression severity, age, and education revealed a striking asymmetry (**Figure 3B**): the apathy–anxiety association was abolished entirely (partial ρ = −0.01, p = 0.634), whereas the apathy–depression association remained robust (partial ρ = 0.37, p < 0.001). This pattern was consistent across all Pedersen apathy subscales and was independent of antidepressant use (**Figure 3B**).

**Figure 3:**
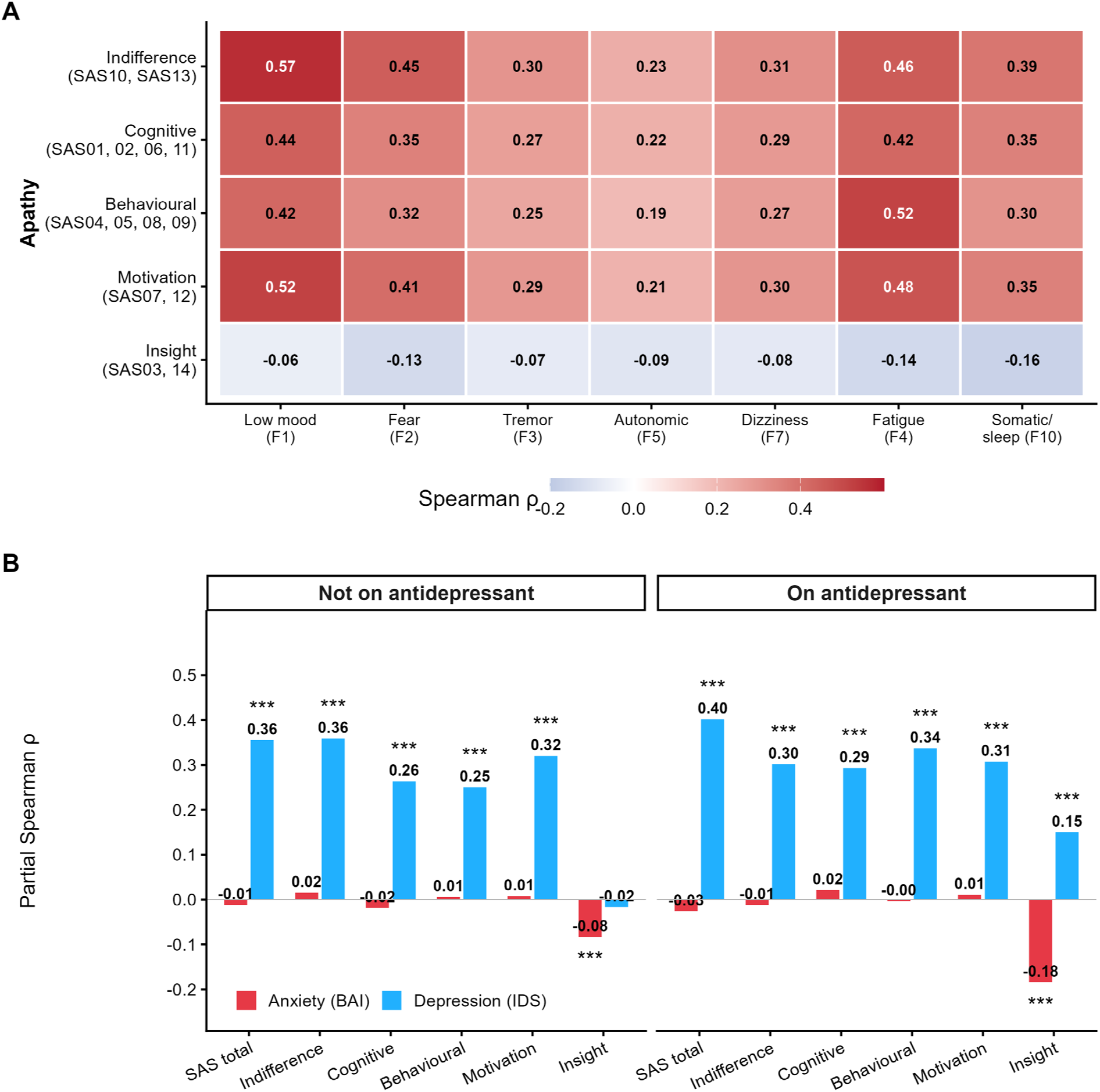
Conditional dependencies between apathy, depression, and anxiety. Pooled NESDO baseline + NESDA Wave E (N = 2,475). (A) Zero-order Spearman correlations between apathy domains and depression/anxiety factors. (B) Partial Spearman ρ between apathy (SAS total and Pedersen subdomains) and depression (IDS-SR) or anxiety (BAI), controlling for age, education, and the other scale, stratified by antidepressant use. The apathy–anxiety correlation drops to near zero when depression is controlled (partial ρ = −0.01, p = 0.634), whereas the apathy–depression association remains significant after controlling for anxiety (partial ρ = 0.37, p < 0.001).

A regularised partial correlation network further visualised this topology, revealing that apathy connects to depression primarily through the “indifference and inertia” node (F6), with no direct edges linking any apathy node to any anxiety node (**Supplementary Figure S4**). The sole exception was Insight: metacognitive awareness of one’s own motivational state showed a weak but significant negative partial correlation with anxiety (partial ρ = −0.10 to −0.14, p < 0.05), suggesting that individuals with greater insight into their apathy also report higher anxiety.

One potential confound in interpreting the apathy–depression overlap is psychomotor retardation, a depressive symptom that may be confounded with behavioural apathy (lack of initiative). In the pooled sample (N = 472), self-reported motor slowing (IDS item 23) correlated most strongly with depressed mood (F1; partial ρ = 0.59) and fatigue (F4; partial ρ = 0.44), but only modestly with apathy factors (F6: ρ = 0.39; F8: ρ = 0.22; F9: ρ = 0.26; **Supplementary Table S2**). This confirms that psychomotor retardation aligns with depressive and somatic dimensions rather than with the motivational deficits captured by the apathy factors.

Moving beyond the inter-relationships between apathy, depression and anxiety, we asked whether these constructs share identical risk factor profiles, or whether the derived factors are driven by different external variables. Separate regression models for each factor score revealed sharply divergent predictor profiles (**Figure 4)**. Female sex was a strong protective factor for apathy, particularly for indifference (F6: β = −0.18, p < .001) and behavioural apathy (F9: β = −0.07, p = .009) while showing no significant association with most anxiety factors. Education was the dominant predictor of cognitive apathy (F8: β = −0.17, p < .001), significantly larger than for any other factor (all Z > 4.7, all p < .001; Z-test for independent coefficients), consistent with cognitive reserve protecting against this specific apathy dimension.

**Figure 4:**
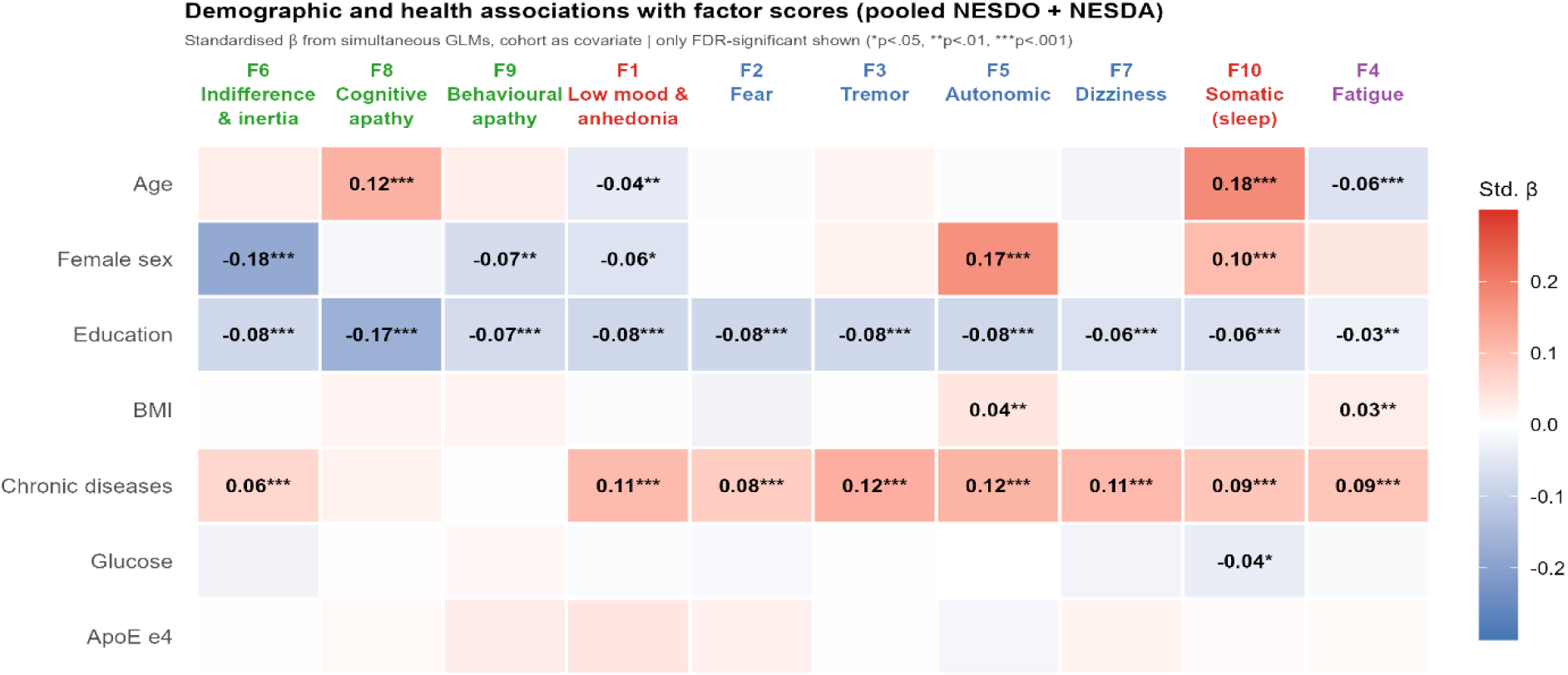
Predictor profiles for 10 symptom factors. Standardised regression coefficients from GLMs predicting each factor score from demographic, clinical, and biological covariates in the pooled sample. FDR-corrected significance: *** p < .001, ** p < .01, * p < .05. Apathy factors show predictor profiles distinct from depression and anxiety.

Crucially, in the NESDO subsample where cognitive data were available (N = 465, age 60–93), global cognition (MMSE) was not a significant predictor of apathy in regression models controlling for age, sex, education, and depression severity (linear: β = −0.08, p = .530; logistic: OR = 1.01, p = .889; **Supplementary Figure S1C–D**). This confirms that the distinct symptom profile of apathy in older adults cannot be merely explained as being an artefact of cognitive decline.

As a secondary clinical-utility analysis, we also tested whether apathy could be reconstructed from standard depression and anxiety items alone. Cross-cohort prediction showed that these measures captured only a minority of individual variation in apathy severity, supporting the need for dedicated apathy assessment rather than inference from mood scales alone (Supplementary Results “Depression and anxiety measures are insufficient to capture clinical apathy”; Supplementary Figure S5).

### Cross-lagged models reveal asymmetric temporal precedence

The cross-sectional analyses above established that apathy is structurally and correlationally separable from depression. A critical question remains: does one precede the other over time? If apathy were simply a downstream consequence of depression, changes in depression should precede changes in apathy, not the reverse. This question necessarily relied on the three-wave longitudinal design of the NESDO cohort, which included repeated apathy assessments at baseline, 2-year and 6-year follow-up (N=292 with complete data). Although smaller than the pooled baseline sample, this longitudinal subsample is the appropriate analytic sample for testing temporal precedence using cross-lagged panel models.

To formally test temporal precedence, we estimated bivariate cross-lagged panel models (CLPMs) between apathy and three symptom dimensions across both time periods (**Figure 5**). Apathy was modelled using the SAS total score because the SAS is a dedicated apathy instrument and the total score captures the overall syndrome-level construct most relevant to clinical prediction. The three symptom dimensions were derived from item-level IDS-SR and BAI data to isolate specific constructs. Anhedonia was indexed by IDS item 21(capacity for pleasure); see Methods-Anhedonia for the rationale. Depressed mood was indexed by three affective items (IDS items 5, 10, 18: feeling sad, mood quality, suicidal ideation), chosen to dissociate it from the somatic, motivational and cognitive items that load onto separate factors. Anxiety was indexed by nine BAI fear and worry items corresponding to EFA factor F2 (items 4, 5, 9–11, 14–17). Somatic BAI items (tremor, dizziness, autonomic symptoms) were excluded from the anxiety composite because they overlap with medical conditions common in older adults and would inflate anxiety–apathy associations.

**Figure 5:**
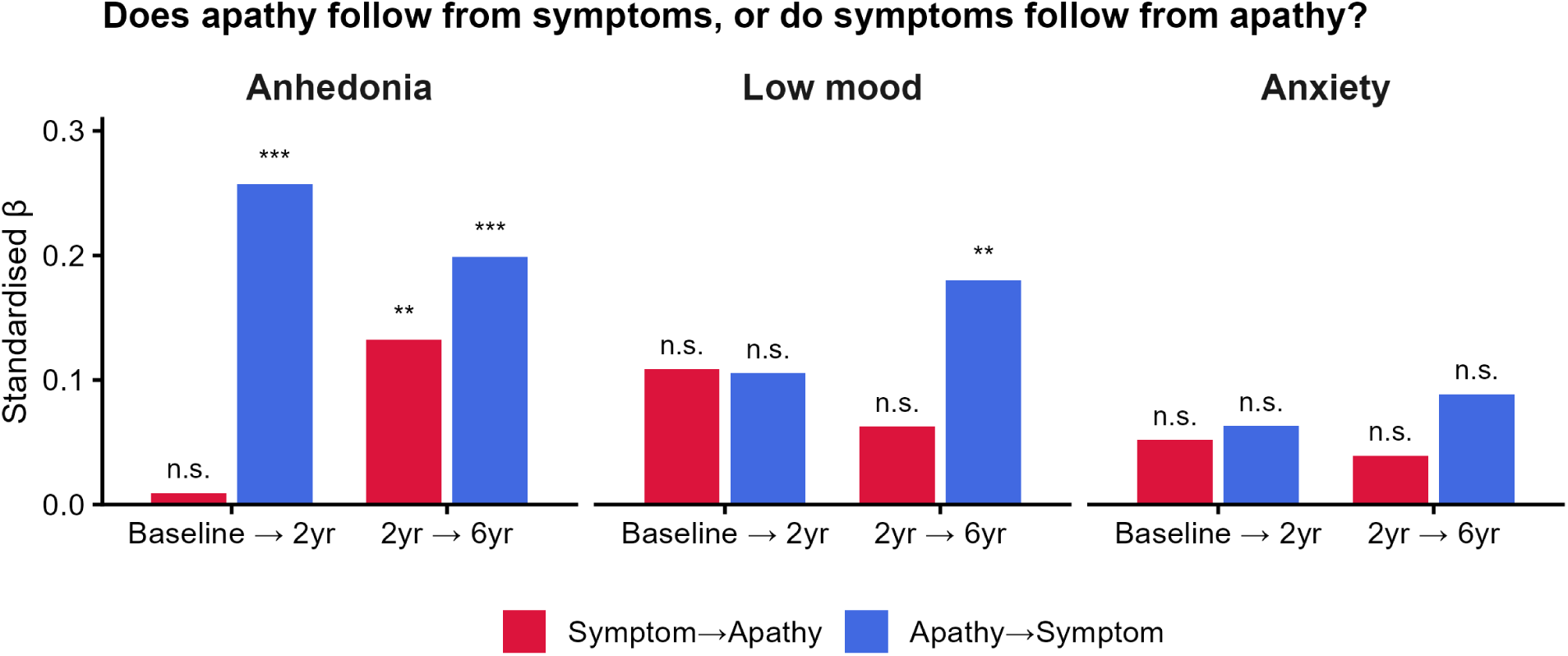
Cross-lagged panel models: apathy and symptom dimensions over 6 years. Standardised cross-lagged coefficients from bivariate CLPMs in NESDO across two periods (baseline → 2-year, 2-year → 6-year) (N=272 for all three waves). Apathy (SAS total) was modelled against anhedonia (IDS capacity for pleasure), depressed mood (IDS composite), and anxiety (BAI fear/worry items, F2). Apathy strongly predicted later anhedonia in both periods (β = 0.26, p < .001; β = 0.20, p < .001); no symptom dimension consistently predicted later apathy.

Apathy strongly predicted subsequent anhedonia in both the baseline-to-2-year period (beta = 0.257, p < 0.001) and the 2-to-6-year period (β = 0.199, p < 0.001), with both effects surviving stringent Bonferroni correction (alpha = 0.004). The reverse pathway (i.e., anhedonia predicting later apathy) was significant only in the second period (β = 0.132, p = 0.008) and did not survive multiple comparison correction. Apathy also predicted later depressed mood in the second period (β = 0.180, p = 0.001), with no significant reverse path observed. No cross-lagged paths emerged between apathy and anxiety in either direction.

Crucially, a comprehensive 4-variable CLPM (modelling apathy, anhedonia, depressed mood, and anxiety simultaneously) confirmed that no pathway from any symptom dimension to later apathy reached significance. Full model parameters and latent growth curve model (LGCM) results are reported in Supplementary Tables S3 and S4. All cross-lagged paths reported above were robust to adjustment for baseline age (apathy → anhedonia T1→T2: β = 0.256; T2→T3: β = 0.224; apathy → sadness T2→T3: β = 0.186; all p ≤ 0.01).The strict asymmetry of these cross-lagged paths confirms that the association between apathy and affective distress does not merely reflect a stable, between-subjects trait correlation—which would produce symmetric pathways—but rather a directional temporal sequence within the disease trajectory.

### Baseline predictors of long-term apathy progression

The CLPMs established that within-person fluctuations in core affective symptoms do not drive subsequent apathy. We next asked whether baseline between-person differences in cumulative clinical burden predict long-term worsening. Worsening overall apathy (residualised 6-year change) was significantly predicted by total baseline depression severity (partial ρ = 0.144), anxiety burden (partial ρ = 0.176), and functional disability, as measured by the WHO-DAS mobility score (partial ρ = 0.204, all p values < 0.05 after multiple comparison correction).

The factor-level analysis showed a more nuanced relationship. The worsening of indifference and inertia (F6) was associated with baseline mood disturbance, whereas the progressive decline in goal-directed behaviour (F9) was entirely independent of mood. Cognitive apathy was the only domain here that was significantly protected by education (Figure 6).

**Figure 6:**
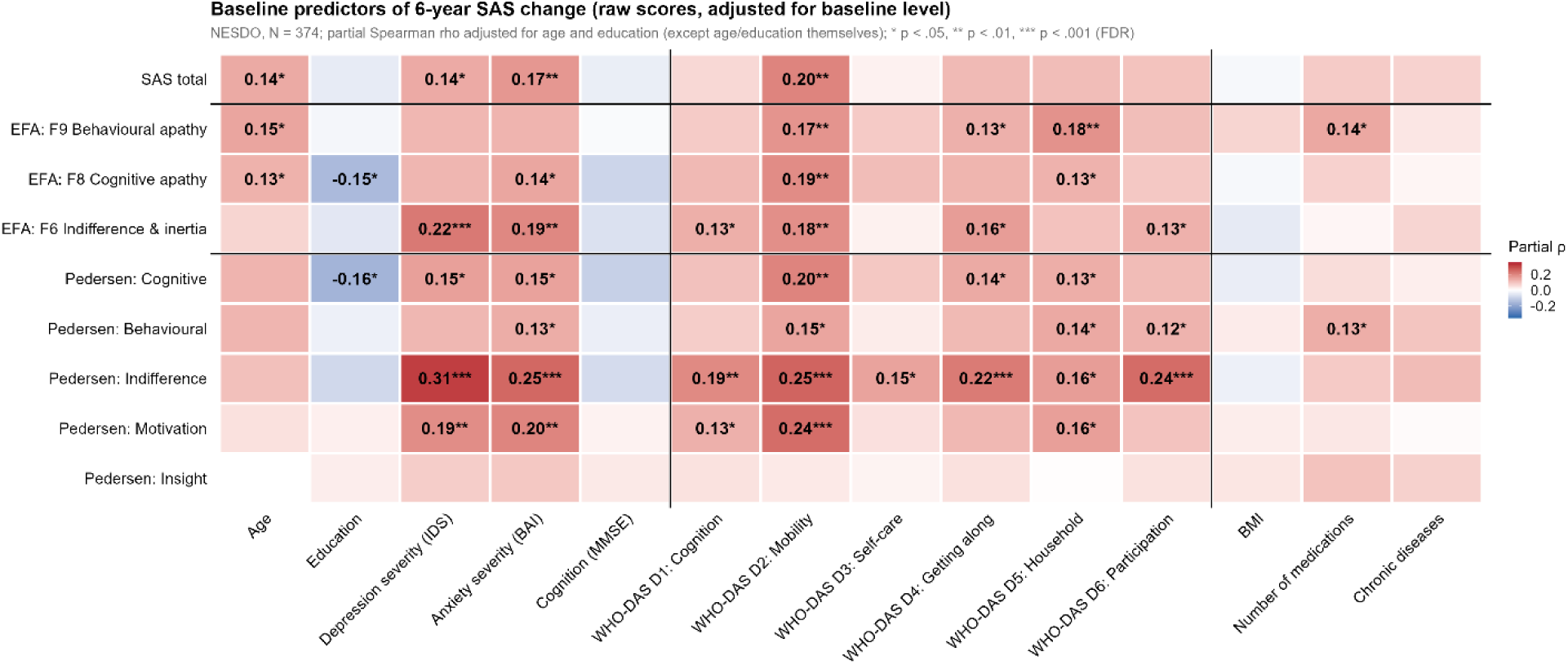
Baseline predictors of 6-year apathy change. Partial Spearman ρ (controlling for age and education) between baseline predictors and residualised change in SAS total and factor scores over 6 years in NESDO (N = 374). Change scores residualised on baseline to control for regression to the mean. The sample size here (N = 374) is larger than that of the cross-lagged panel models in Figure 5 (N = 292) because the present analysis requires data at only two waves (baseline and 6-year follow-up), whereas the CLPMs require complete data at all three NESDO waves.* FDR p < .05, ** p < .01.

### APOE e4 genotype does not drive apathy in psychiatric cohorts

Finally, we explored whether this longitudinal decline might be driven by underlying neurodegenerative processes. Because apathy is highly prevalent in Alzheimer’s disease and frequently serves as an early neurobehavioural prodrome, we tested whether the Apolipoprotein E (APOE) e4 allele predicts apathy trajectories. We hypothesised that if apathy reflects preclinical neurodegeneration, e4 carriers should exhibit significantly higher severity and faster longitudinal worsening.

APOE genotype data were available for 2,451 NESDA and 487 NESDO participants. The ε4 allele frequency was 16.6% in NESDA and 16.0% in NESDO, yielding carrier prevalences of 30.8% and 30.2% respectively—consistent with expected rates for Northern European populations.^23,24^ Genotype distributions were comparable across cohorts, and Hardy-Weinberg equilibrium was confirmed (both p > 0.05). Heterozygous and homozygous carriers were pooled for all analyses (total homozygous n = 69), and the e2 allele showed no association with apathy in any model.

In the pooled sample, e4 carrier status was not significantly associated with cross-sectional apathy after adjusting for age, sex, education, depression severity, and cohort. Interaction models (e4 × age, sex, or depression) yielded strictly null results. Although females over 75 years old showed numerically higher apathy prevalence among carriers (82.6% vs 65.7%, **Figure 7A**), this involved only 23 carriers and did not approach significance (p = 0.187).

**Figure 7:**
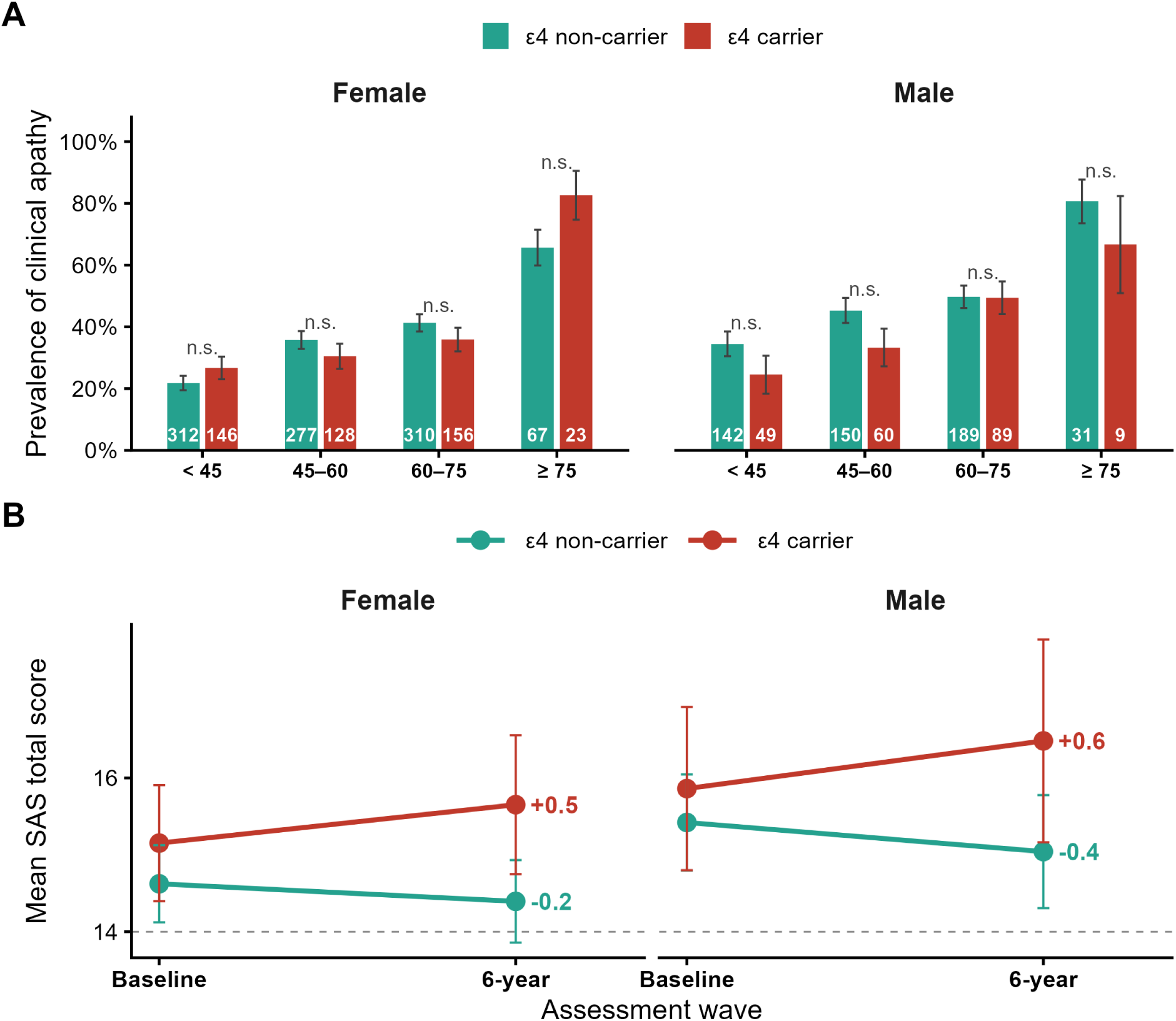
APOE ε4 genotype and apathy. (A) Prevalence of clinical apathy (SAS ≥ 14) by age group and ε4 carrier status, stratified by sex. (B) Longitudinal apathy trajectories by ε4 carrier status and sex over 6 years in NESDO.

Longitudinally, ε4 carriers showed a non-significant trend toward increasing apathy over 6 years (female: +0.5, male: +0.6) while non-carriers declined (female: −0.2, male: −0.4; **Figure 7B**), but a linear mixed model confirmed the time × e4 interaction did not reach significance (β = 0.198, p = 0.057) and was further attenuated after adjusting for time-varying depression (β = 0.140, p = 0.151). Taken together, genetic neurodegenerative risk does not exert a meaningful independent effect on apathy in these psychiatric cohorts.

## Discussion

In this study of 2,482 adults with depressive or anxiety disorders, we demonstrate that apathy is structurally and correlationally separable from depression and anxiety across the adult lifespan. Within the longitudinal NESDO subsample (N=292 with complete three-wave item-level data spanning six years), cross-lagged panel models further reveal an asymmetric temporal sequence: early motivational deficits strongly predict the subsequent worsening of depressed mood and anhedonia. Crucially, no symptom dimension consistently predicted later apathy. If this relationship merely reflected a stable between-subjects correlation—whereby more severely depressed individuals concurrently exhibit greater apathy—the longitudinal cross-lagged paths would be symmetric. Instead, this strictly asymmetric trajectory demonstrates that motivational withdrawal is not merely a downstream residue of chronic depression. It potentially functions as an upstream driver of affective decline.

This temporal asymmetry is further supported by the structural independence of these constructs. Over 60% of older adults and 30% of younger adults with mood disorders met the clinical apathy threshold, and the overlap with depression was substantial. Yet, item-level factor analysis across 62 symptoms consistently separated multidimensional apathy—indifference, cognitive apathy, and behavioural apathy—from anhedonia, depressed mood, somatic fatigue, and anxious arousal. This structure replicated across an independent cohort and remained stable over six years of measurement invariance testing. This confirms genuinely distinct clinical dimensions rather than sample-specific artefacts. Partial correlations demonstrated that the apathy–anxiety association was abolished entirely after controlling for depression, whereas the apathy–depression link remained robust. This demonstrates that apathy is selectively coupled to depression, rather than being a generic symptom of overall affective distress.

Crucially, this specific coupling is highly stable across the lifespan. Although advancing age and depression both independently predict greater apathy, their effects are strictly additive. The depression-by-age interaction was completely null, meaning the gap in apathy severity between depressed and non-depressed individuals remains constant from early adulthood into old age. This enduring link reflects a stable coupling between motivation and mood, two overlapping but structurally independent constructs. The relationship is not explained by age-related cognitive decline or psychomotor retardation.

These findings converge with a growing body of translational neuroscience. Preclinical models and human neuroimaging consistently demonstrate that the neural circuits governing effort allocation (‘wanting’) are biologically dissociable from the hedonic hotspots governing consummatory pleasure (‘liking’),^1,9,16^ with motivation relying heavily on dopaminergic projections within the frontostriatal network and anterior cingulate cortex.^10,17^ Our data provide indirect but converging clinical evidence for this dissociation. Cross-sectionally, apathy showed a markedly different predictor profile from depression: physical comorbidity was strongly associated with depressive and anxiety factors but only weakly or not at all with apathy (**Figure 4**).

Yet longitudinally, a different picture emerged. Mobility disability and anxiety severity were the strongest predictors of apathy worsening over 6 years, whilst cognitive decline (MMSE) and APOE ε4 genotype were null. This dissociation between concurrent and prospective predictors indicates that apathy does not simply co-occur with physical decline at a given moment, but that sustained exposure to functional limitation gradually erodes motivational capacity.

This interpretation aligns with preclinical evidence that chronic stress progressively downregulates mesolimbic dopamine signalling, degrading effort-based decision-making before hedonic processing is compromised.^18^ The implication is that motivational circuits are not damaged by depression per se, but by the broader physiological cost of living with chronic illness, a cost that disability and affective distress contribute to independently. Patients do not cease goal-directed behaviour because they have lost the capacity for pleasure. The deterioration of effort allocation comes first, initiating a cascade of behavioural withdrawal that ultimately precipitates anhedonia and depressed mood.

Complicating this clinical picture, standard pharmacological treatments frequently fail to resolve motivational deficits and may compound them. Non-SSRI antidepressants (including SNRIs such as venlafaxine and duloxetine, and noradrenergic agents such as mirtazapine) and benzodiazepines were independently associated with increased apathy severity in our regression models, even after controlling for depression and anxiety. We do not claim that these medications directly cause apathy. Confounding by indication is substantial in observational cohorts: patients prescribed non-SSRI agents have typically failed first-line SSRI treatment, reflecting more treatment-resistant illness. Nevertheless, these associations converge with growing evidence that serotonergic treatments fail to rescue motivation and may induce iatrogenic apathy through blunting of reward-related neural activity.^11,14,15,25–27^

In preclinical models, serotonin transporter inhibitors are ineffective at restoring effort-based choice, whereas dopamine transporter inhibitors successfully rescue motivated behaviour. This pharmacological dissociation mirrors the clinical pattern we observe here.^28^ While SSRI use has been consistently linked to greater apathy in Parkinson’s disease,^29,30^ our findings extend this concern to primary psychiatric populations. Regardless of the causal direction, the clinical imperative is clear: apathy must be systematically quantified and monitored throughout pharmacological management of depression, both as a core outcome measure and as a potential treatment-emergent adverse effect.

A major barrier to addressing this therapeutic gap is that apathy is rarely quantified specifically during routine psychiatric diagnosis or depression management. Unfortunately, this oversight cannot be bypassed by simply re-analysing established depression and anxiety instruments. Our cross-cohort prediction models revealed that standard depression and anxiety scales miss up to 88% of the individual variance in apathy severity. Even the most informative items,which were themselves apathy-adjacent, achieved only modest group-level discrimination and poor individual-level prediction. Consequently, clinical trials relying exclusively on broad affective measures are fundamentally unequipped to detect motivational deficits. Dedicated apathy instruments must be incorporated into both routine care and future intervention trials. To our knowledge, this study represents the largest longitudinal psychiatric dataset to employ a dedicated, item-level apathy questionnaire. We hope that this work encourages ongoing mega-cohorts and future study planners to include validated apathy assessments as standard protocol.

Realising this ambition, however, requires acknowledging the boundaries of the present evidence. Several methodological constraints temper our conclusions. First, traditional cross-lagged panel models establish temporal precedence but inherently conflate between-person trait differences with within-person temporal dynamics. A definitive test isolating the strictly within-individual sequence would require a random-intercept CLPM, which was precluded here by the sample size and the three-wave limit of the longitudinal cohort. However, our complementary analyses provide converging evidence for the within-person trajectory. Latent growth curve models confirmed that baseline depression predicts concurrent apathy but not its rate of change, whilst residualised change analyses approximated within-person worsening by partialling out baseline variance. Both approaches converge to suggest that mood does not drive within-person apathy progression. Nonetheless, observational data cannot definitively prove mechanistic causality; randomised experimental manipulation of apathy would be required to confirm the true causal direction.

Second, anhedonia was operationalised using a single IDS-SR item (capacity for pleasure) rather than a multidimensional instrument, and this warrants careful interpretation. The present cohort did not include a specific anhedonia questionnaire, so we cannot disaggregate anticipatory from consummatory pleasure, nor social from physical reward, nor motivational from hedonic components. We acknowledge that this single-item proxy is a coarse instrument and cannot resolve the substructure of anhedonia (e.g., anticipatory vs consummatory pleasure). At the same time, diminished capacity for pleasure is widely regarded as a core feature of anhedonia in DSM-5 and ICD-11, which provides some reassurance that the item indexes the construct’s central phenomenology even if it cannot resolve its substructure. However, item-level analyses show this single-item indicator is empirically distinct from the SAS (**Supplementary Figure S7**), and a 2-item anhedonia composite (see Methods – Anhedonia) did not change the cross-lagged estimates (**Supplementary Tables S5 and S6**). Third, the Starkstein Apathy Scale (SAS), whilst well-validated, was originally developed for neurological populations. More modern instruments, such as the Apathy Motivation Index,^31^ the Lille Apathy Rating Scale,^32^ and the Dimensional Apathy Scale,^33^ offer finer-grained measurement of motivational subdomains. In particular, the "indifference and inertia" factor (F6) in our analysis likely conflates environmental indifference with a lack of self-initiated behaviour. Instruments like the AMI and LARS more explicitly separate emotional apathy—reduced concern or empathy for others—from behavioural initiation deficits. Previous work has shown that emotional apathy is not positively associated with, and may even be negatively related to, depression,^6,31^ a distinction that could not be fully resolved with the SAS and warrants dedicated investigation. Fourth, the APOE analysis was underpowered in the oldest age band (≥75 years: n = 130, of which only 32 were ε4 carriers), precisely where neurodegenerative contributions are most plausible. Survival bias may further attenuate the true ε4 effect, and future studies should adjust for cognitive status to definitively separate neurodegenerative from affective apathy. Fifth, whilst the combined sample spans ages 24 to 93, participants are drawn from a predominantly Northern European demographic, and generalisability to other populations remains to be established.

The clinical imperative is nonetheless clear. By establishing apathy as a structurally independent, temporally antecedent predictor of depressive worsening, these findings justify the early identification and targeted treatment of motivational deficits—before they cascade into the full syndrome of major depression. The development of circuit-directed interventions, including dopaminergic agents, focused behavioural activation, or neuromodulation designed to rescue effort-based decision-making, represents a critical and currently underexploited therapeutic frontier.

## Methods

### Ethics statement

Data were drawn from two Dutch longitudinal cohort studies of depression and anxiety: the Netherlands Study of Depression and Anxiety (NESDA) and the Netherlands Study of Depression in Older Persons (NESDO).^19–21^ NESDA was approved by the Medical Ethical Review Board (METC) of each participating centre in Amsterdam, Leiden, and Groningen (reference: METC 2003-183). NESDO was approved centrally by the Ethical Review Board of the VU University Medical Centre, and subsequently by the local review boards of Leiden University Medical Centre, University Medical Centre Groningen, and Radboud University Medical Centre Nijmegen. All participants in both studies provided written informed consent in accordance with the Declaration of Helsinki.

### Participants

NESDA is a multisite longitudinal cohort study designed to examine the long-term course and consequences of depressive and anxiety disorders.^19,20^ Participants (N = 2,981 at baseline; age 18–65 years) were recruited from community, primary care, and specialised mental health settings across the Netherlands between 2004 and 2007. Follow-up assessments were conducted at 1, 2, 4, 6, 9, and 12 years post-baseline follow-up (assessed approximately 2010-2013), which overlaps with the NESDO baseline (2007-2010). This was also the first NESDA wave at which the Starkstein Apathy Scale was administered, which is why we used it as the cross-sectional anchor. Full details of the NESDA design and procedures have been published previously.

NESDO is a prospective cohort study of depression in older adults (age >= 60 years).^21^ Participants (N = 510 at baseline; age 60–93 years) were recruited from mental health and primary care settings between 2007 and 2010. Follow-up assessments were conducted at 2 years (N = 299; 58.6% retention) and 6 years (N = 401; 78.6% retention), with 292 who completed all three waves Full details of the NESDO design have been published previously.^21^

The present analysis pooled NESDA Wave E (6-year follow-up; N = 2,256) with NESDO baseline (N = 510), yielding a combined sample of N = 2,766 with apathy data spanning ages 24 to 93 years at baseline. After exclusions for missing data on key variables, the effective sample sizes for individual analyses were as follows: prevalence estimates N = 2,482, regression models N = 2,469, comorbidity analyses N = 2,475, EFA N = 510, CFA N = 2,256, CLPMs N = 292, and network analysis N = 2,460. The demographics and clinical characteristics of both cohorts are summarised in **Table 1**.

### Measures

#### Apathy

Apathy was assessed using the 14-item Starkstein Apathy Scale (SAS), a self-report measure originally developed in Parkinson’s disease.^34^ Items are scored 0–3 (range 0–42), with higher scores indicating greater apathy. Clinical apathy was defined as SAS ≥ 14, following established thresholds.^34^ Item-level data were available at all three waves in NESDO and at Wave E in NESDA. Internal consistency was good in both cohorts (Cronbach’s α = 0.80 in NESDO baseline, n = 449; α = 0.80 in NESDA Wave E, n = 2,074).

#### Depression

Depressive symptoms were measured using the 28-item Inventory of Depressive Symptomatology — Self Report (IDS-SR).^35^ Total scores range from 0 to 84, with severity categories defined as: none (0–13), mild (14–25), moderate (26–38), severe (39–48), and very severe (>= 49). In analyses stratifying by depression status, depression was defined as IDS-SR total >= 14 (i.e., mild severity or above). Item-level data (items 1–28, excluding item 9 which assesses diurnal variation direction rather than severity) were used in the factor analyses. Following standard IDS-SR scoring, items 11/12 (appetite decrease/increase) and items 13/14 (weight decrease/increase) are distributed by the cohorts as single collapsed severity scores (0–3) reflecting whichever direction the respondent endorsed; we used these direction-agnostic severity items in all factor and network analyses. IDS item 23 (self-reported psychomotor slowing) was analysed separately as a marker of psychomotor retardation. Internal consistency was excellent (α = 0.91 in NESDO baseline, n = 433; α = 0.90 in NESDA Wave E, n = 2,060).

#### Anhedonia

Anhedonia was operationalised using a single IDS-SR item: item 21, ‘capacity for pleasure or enjoyment (excluding sex)’. Capacity for pleasure is the core feature of anhedonia in major diagnostic frameworks.^1,16,18,36^ Although a 3-item IDS-SR composite, namely items 19 (*"general interest"*), 21 (*"capacity for pleasure"*) and 22 (*"interest in sex"*), has been used as an anhedonia proxy in previous studies,^37,38^ we did not adopt it here for the conceptual and empirical reasons given below.

*Item 22 (interest in sex)* was excluded because we have previously demonstrated that loss of interest in sex is one of the strongest indicators of *pure depression* (depression in the absence of apathy or anhedonia), with each construct measured using established scales.^6^ In the resulting Apathy–Depression–Anhedonia Measure (ADAM),^6^ reduced libido is grouped with the four depression-defining items (sadness, guilt, crying, reduced libido), not with anhedonia or apathy. Including a depression-specific symptom in the anhedonia construct would conflate the two dimensions the present analyses aim to dissociate.

*Item 19 (general interest)* was excluded because loss of general interest is itself a form of cognitive apathy: the item indexes engagement with people, work and hobbies (motivational and initiation deficits) rather than consummatory pleasure.^39^ Salamone and Correa^16^ further argue that conflating reduced interest or effort with reduced hedonic capacity has obscured the dopaminergic biology of motivation in the depression literature. Indeed, in the original paper validating this anhedonia subscale, item 21 (“capacity for pleasure”) correlated strongly with the Snaith-Hamilton Pleasure Scale (SHAPS), an established measure of anhedonia,^40^ whereas the general interest item was only moderately correlated with SHAPS.^41^ Item-level analyses in the present pooled sample (NESDO baseline + NESDA Wave E; N = 2,609; **Supplementary Figure S7**) confirmed this distinction empirically. Item 19 was significantly more strongly correlated with apathy than item 21 was for SAS total (Steiger Z = +3.13, p = .002, FDR = .005), behavioural apathy (F9: Z = +3.67, p < .001, FDR = .001) and the Pedersen Behavioural subscale (Z = +4.22, p < .001, FDR < .001), whereas item 21 was more strongly correlated than item 19 only with the apathy domain closest to consummatory anhedonia (Pedersen Indifference: Z = −2.54, p = .011, FDR = .025). This pattern is consistent with item 19 indexing motivational engagement (a cognitive- and behavioural-apathy facet) rather than hedonic capacity.

Although a single-item indicator is coarse, IDS21 (‘capacity for pleasure’) is the more conservative choice for the present analysis because it does not import apathy or depression variance into the anhedonia side of the cross-lagged models.

Nevertheless, as a sensitivity check, we re-ran all bivariate and 4-variable CLPMs using a 2-item composite (mean of items 19 and 21); the cross-lagged estimates matched the main 1-item analysis closely (**Supplementary Tables S5 and S6**), so the asymmetric apathy → anhedonia precedence does not depend on the single-item indicator.

#### Anxiety

Anxiety was assessed with the 21-item Beck Anxiety Inventory (BAI).^42^ Total scores range from 0 to 63, with severity categories defined following Kabacoff et al.: normal (0–9), mild (10–18), moderate (19–29), and severe (>= 30).^43^ Clinical anxiety in the comorbidity analysis was defined as BAI total >= 10 (i.e., mild severity or above). Internal consistency was excellent (α = 0.94 in NESDO baseline, n = 453; α = 0.92 in NESDA Wave E, n = 2,084).

#### Education

Education level was recorded as an ordinal variable in both cohorts but using different category schemes: NESDA used a 9-point ordinal scale of highest completed education (1 = primary school to 9 = university plus postdoctoral training; variable aeducat), and NESDO used an 11-point ordinal scale of highest completed diploma (variable Opleidin). To enable pooling across cohorts, education was z-standardised within each cohort separately, preserving rank-order information whilst harmonising scale differences. Years of schooling were available in NESDA but were not used, to maintain comparability with the NESDO categorical scheme.

#### Cognitive function

Global cognitive function was assessed in the NESDO subsample using the Mini-Mental State Examination (MMSE; range 0–30), administered at baseline.

#### Functional disability

Both cohorts administered the 36-item WHO Disability Assessment Schedule 2.0 (WHO-DAS), scored identically using the Dutch manual algorithm to yield a total disability score and six domain scores (cognition, mobility, self-care, getting along, household, and participation).^44^ Internal consistency across the 32 domain items (D1–D6) was excellent (α = 0.95 in NESDO baseline, n = 393; α = 0.96 in NESDA Wave E, n = 2,038).

#### Chronic diseases

The number of chronic somatic diseases was ascertained through a structured self-report interview covering 13 disease categories: chronic non-specific lung disease (COPD), cardiovascular disease (including coronary heart disease, heart failure, and cardiac arrhythmia but excluding isolated angina pectoris), peripheral vascular disease, diabetes mellitus, stroke, arthritis or rheumatism (merged into a single category because patients frequently confuse these conditions), cancer, peptic ulcer, intestinal disease (excluding functional symptoms such as diarrhoea and constipation), liver disease, epilepsy, thyroid disease, and other chronic diseases. Osteoarthritis and rheumatism were counted as one condition. The resulting count variable (range 0–13) was used in all analyses. In a secondary count, hypertension was additionally included; analyses in this paper used the primary count excluding hypertension. In NESDA, the equivalent variable was derived using the same self-report interview format.

#### Medication use

Current psychotropic medication use was ascertained through inspection of medication containers brought by participants to the assessment visit. Each medication was classified using the World Health Organisation Anatomical Therapeutic Chemical (ATC) classification system. Antidepressant use was defined as any medication with the ATC code prefix N06A, which encompasses the following subclasses: selective serotonin reuptake inhibitors (SSRIs; ATC N06AB— e.g., citalopram, sertraline, paroxetine, fluoxetine, fluvoxamine), non-selective monoamine reuptake inhibitors (TCAs; ATC N06AA — e.g., amitriptyline, nortriptyline, clomipramine, imipramine), other antidepressants (ATC N06AX — e.g., venlafaxine [N06AX16], mirtazapine [N06AX11], duloxetine [N06AX21], bupropion [N06AX12], trazodone [N06AX05]), and monoamine oxidase inhibitors (ATC N06AF/N06AG). Benzodiazepine use was defined as any medication with the ATC code prefix N05BA (anxiolytic benzodiazepine derivatives) or N05CD (hypnotic benzodiazepine derivatives). In the main analyses, medication was classified into four binary variables: any antidepressant (N06A, yes/no), SSRI (N06AB, yes/no), non-SSRI antidepressant (N06A but not N06AB, yes/no), and benzodiazepine (N05BA/N05CD, yes/no). Participants using more than one antidepressant from different subclasses were classified according to the broadest category (any antidepressant) or according to each subclass simultaneously. Medication frequency was recorded (infrequent: less than 50% of the time or as needed; frequent: more than 50% of the time or daily); any recorded use was classified as positive.

#### APOE genotyping

Apolipoprotein E (APOE) genotype data were available for 2,451 NESDA and 487 NESDO participants. In NESDA, APOE genotype was derived from two single nucleotide polymorphisms (SNPs): rs429358 and rs7412. The C allele at rs429358 defines the ε4 allele, and the T allele at rs7412 defines the ε2 allele. These two SNPs were used to assign standard APOE genotypes (ε2/ε2, ε2/ε3, ε2/ε4, ε3/ε3, ε3/ε4, ε4/ ε4). Participants without available DNA (n = 530) were excluded from APOE analyses. In NESDO, APOE genotype was extracted from the Genotype column in the baseline dataset, filtering for entries matching the pattern (the column also contained non-APOE SNP data, which were excluded). APOE was operationalised as a dichotomous ε4 carrier status variable (0 = non-carrier; 1 = carrier [ε3/ε4 or ε4/ ε4), with heterozygous and homozygous ε4 carriers pooled. As a sensitivity analysis, an additive allele-dose model (ε4 count: 0, 1, or 2) was also fitted; results were consistent with the dichotomous carrier analysis and are not tabulated separately. The ε2 allele was analysed separately but showed no significant associations. Hardy–Weinberg equilibrium was confirmed in both cohorts (both p > 0.05).

#### Missing values

In both cohorts, derived scale scores that could not be computed due to excessive item-level missingness were coded as -1; system-missing values were coded as -3. These sentinel values were recoded to NA prior to analysis. Participants with missing data on key outcome variables were excluded from the relevant analysis.

### Statistical analysis

All analyses were conducted in R (version 4.5.2) using the following packages: psych (v2.4.12; EFA and parallel analysis), lavaan (v0.6-19; CFA, measurement invariance, structural equation modelling), GPArotation (v2024.3-1; oblimin rotation), ppcor (v1.1; partial correlations), estimatr (v1.0.4; cluster-robust standard errors), bootnet (v1.6; network estimation) and qgraph (v1.9.8; network visualisation), glmnet (v4.1-8; elastic net regularisation), and tidyverse (v2.0.0; data processing and visualisation). All p-values from regression-based analyses were corrected for multiple comparisons using the Benjamini–Hochberg false discovery rate (FDR) procedure unless otherwise stated. Variance inflation factors (VIF) were checked for multicollinearity in all regression models (threshold > 5). Residual normality was assessed via Shapiro–Wilk tests where applicable.

### Cross-sectional associations

Associations between apathy, age, and depression were examined using linear regression (continuous SAS) and logistic regression (clinical apathy, SAS >= 14) in the pooled NESDA + NESDO sample (N = 2,482 for linear; N = 2,469 for logistic). Running means with a 5-year sliding window (+/-2.5 years; minimum 10 observations per window) were used to visualise age trends (Figure 1A; Supplementary Figure S1A). An age x depression interaction term tested whether the depression–apathy association varied across the lifespan (Table 2). A sensitivity analysis in the NESDO subsample (N = 465) added MMSE as a covariate to assess whether the age–apathy association was confounded by cognitive decline (Supplementary Figure S1C–D).

### Exploratory factor analysis

EFA was performed on the NESDO baseline sample (N = 510) using all 62 symptom items (14 SAS + 21 BAI + 27 IDS-SR). We deliberately conducted the EFA in NESDO (N = 510) rather than the larger NESDA Wave E sample (N = 2,256) for three reasons. First, the question of structural separability is most clinically pressing in older adults, where apathy is most prevalent and most often conflated with depression; establishing structure in this population and then testing generalisation to a younger, larger cohort is the stronger inferential direction. Second, N = 510 is well within recommended ranges for EFA with items of this communality,^45^ and sampling adequacy was empirically excellent (KMO = 0.944). Third, reserving the larger NESDA sample for CFA maximises statistical power for the confirmation step, where large N is most valuable for stable fit-index estimation. avoiding the reduced generalisability that can arise when EFA is run in very large samples, where statistically detectable but substantively trivial factors may be extracted. Sampling adequacy was assessed using the Kaiser–Meyer–Olkin (KMO) measure and Bartlett’s test of sphericity.

The number of factors was determined by Horn’s parallel analysis (1,000 simulated datasets; factor analysis extraction). Factors were extracted using maximum likelihood estimation with oblimin rotation (allowing correlated factors). Items with loadings >= 0.30 on their primary factor were retained for the confirmatory model. Factor scores for all subsequent analyses were computed as the unweighted mean of items assigned to each factor, providing interpretability and replicability across samples at the cost of ignoring differential item weightings. Full item-level loadings and communalities are reported in Supplementary Table S1; the inter-factor correlation matrix is shown in Supplementary Figure S3. All 62 items (14 SAS, 21 BAI, 27 IDS-SR) share a common four-point Likert response format scored 0–3, so items entered the EFA on a harmonised scale without dichotomisation or rescaling. This minimises the risk that factors are driven by differences in response formats across instruments.

### Confirmatory factor analysis

The EFA-derived 10-factor model was tested via CFA in three independent samples: NESDO 2-year follow-up (assessed 2009–2012; N = 299), NESDO 6-year follow-up (assessed 2013–2016; N = 401), and NESDA Wave E (6-year follow-up, assessed 2010–2013; N = 2,256). All CFA models used maximum likelihood estimation with full information maximum likelihood (FIML) for missing data and standardised latent variables (std.lv = TRUE). Model fit was evaluated using CFI, TLI, RMSEA, and SRMR. We note that conventional CFI/TLI thresholds (>= 0.90) were developed for simpler models and are systematically penalised in complex multi-factor models with many items; RMSEA (<= 0.08 acceptable, <= 0.06 good) and SRMR (<= 0.10) are therefore prioritised in fit evaluation. Nested model comparisons used chi-square difference tests. As a convergent validity check, an independent EFA on the NESDA Wave E data was also conducted to verify that a comparable factor structure emerged in a different cohort. When using standardised latent variables (std.lv = TRUE), CFA solutions are subject to sign indeterminacy, i.e.: the direction of a latent variable is arbitrary if all loadings are reflected. In the NESDO CFA, F10 (IDS+BAI Somatic/sleep) yielded all-negative loadings owing to this indeterminacy. To resolve this, an automatic sign-flipping correction was applied: when the majority of a factor’s standardised loadings were negative, all loadings and inter-factor correlations involving that factor were reflected. This correction does not affect model fit, variance explained, or substantive interpretation; it merely ensures consistent sign conventions across samples.

### Measurement invariance

Longitudinal measurement invariance was tested across the three NESDO waves (baseline, 2-year, 6-year) using a simplified 10-factor CFA model (top 3–5 highest-loading items per factor; 26 items). The full 62-item model exceeded estimation capacity with three-wave longitudinal data at this sample size; the simplified model retained items with the strongest factor loadings and reproduced the full model’s factor structure. Three levels of invariance were tested sequentially: configural (equal factor structure across waves), metric (equal factor loadings), and scalar (equal intercepts). Invariance at each level was supported if DELTA-CFI > -0.01 and DELTA-RMSEA < 0.015, following Chen (2007). Latent means were extracted from the scalar invariance model with baseline as the reference group (Table 3).

### Factor profiling

To identify distinct predictor profiles for each of the 10 factors, separate linear regression models were fitted in the pooled sample predicting each factor score from demographic, clinical, and health covariates: age, sex, education (within-cohort z-score), BMI, fasting glucose, APOE ε4 carrier status, chronic disease count, and cohort (Figure 4). All continuous predictors were z-standardised. VIF was checked for multicollinearity (threshold > 5). Antidepressant use was not included as a predictor to avoid collider bias (i.e., antidepressant use is influenced by depression severity, which in turn predicts apathy; conditioning on a collider can introduce spurious associations). Residual normality was assessed via Shapiro–Wilk tests.

### Partial correlation and network analysis

Zero-order and partial Spearman correlations were computed between apathy subdomains (both EFA-derived factors and Pedersen et al. subscales) and depression (IDS-SR) and anxiety (BAI), controlling for age, education, and the other scale (Figure 3). Analyses were stratified by antidepressant use to assess whether medication status confounded the observed associations. A Gaussian graphical model (partial correlation network) was estimated on the pooled sample (N = 2,460 complete cases) using the EBICglasso algorithm with the extended Bayesian information criterion (EBIC; gamma = 0.5) for regularisation (Supplementary Figure S4). Fourteen nodes were included: 10 EFA factor scores and 4 demographic/health variables (age, education, BMI, chronic disease count). Node centrality was quantified using strength, betweenness, and closeness indices. Stability of centrality estimates was assessed using case-dropping bootstrap (500 iterations), with the CS-coefficient (proportion of cases that can be dropped while maintaining a correlation >= 0.7 with the original centrality ordering) as the stability metric (CS >= 0.25 acceptable, >= 0.50 preferred). Edge accuracy was assessed using nonparametric bootstrap (1,000 iterations).

### Cross-lagged panel models

Temporal precedence between apathy and specific symptom dimensions was tested using bivariate cross-lagged panel models (CLPMs) across the three NESDO waves (N = 292 with data at all three waves). Three separate CLPMs examined apathy (SAS total) in relation to: (i) anhedonia (IDS item 21: capacity for pleasure), (ii) depressed mood (IDS items 5, 10, 18: feeling sad, mood quality, suicidal ideation), and (iii) anxiety (BAI anxiety/worry subscale corresponding to EFA factor F2: nine items — BAI 4, 5, 9–11, 14–17). Somatic BAI items (tremor, dizziness, autonomic symptoms) were excluded from the anxiety composite because they overlap with medical conditions common in older adults and would inflate anxiety–apathy associations.

All variables were z-standardised within each wave (using wave-specific means and standard deviations in wide format), thereby preserving cross-lagged path interpretation while removing mean-level differences between waves. Models were estimated with FIML and included autoregressive paths, cross-lagged paths, and concurrent correlations. A full 4-variable CLPM (apathy, anhedonia, depressed mood, and anxiety simultaneously) was fitted as a sensitivity analysis (Supplementary Figure S6). Cross-lagged paths surviving Bonferroni correction (alpha = 0.05/12 for 12 paths across 3 bivariate models) are highlighted in Figure 5. Full model parameters are reported in Supplementary Table S3.

A complementary latent growth curve model (LGCM) was fitted to test whether baseline symptom levels predicted the rate of apathy change over 6 years. Baseline anxiety (BAI F2), anhedonia (IDS item 21), depressed mood (IDS composite of items 5, 10, 18), and MMSE were entered as predictors of the latent intercept and slope of apathy (Supplementary Table S4).

In a sensitivity analysis, baseline age was added as a time-invariant covariate predicting each variable at every wave; the headline cross-lagged paths were unchanged in direction, magnitude, or significance.

### Change correlates

Predictors of 6-year apathy change were examined in the NESDO longitudinal sample (N = 377 with data at both baseline and 6-year follow-up). Change scores (6-year minus baseline z-score) were computed for SAS total and the three apathy factor scores (F6: Indifference and inertia, F8: Cognitive apathy, F9: Behavioural apathy). To address regression to the mean, each change score was residualised on its baseline value. Spearman correlations (partial, controlling for age and education) were computed between baseline predictors and residualised change scores, with FDR correction (Figure 6).

### Cross-cohort prediction

To quantify the shared variance between apathy and depression/anxiety symptoms, an elastic net prediction model was trained on NESDO baseline (N = 388) and externally validated on NESDA Wave E (N = 1,835). Feature selection used minimum redundancy–maximum relevance (mRMR) ranking with a k-sweep (k = 1 to 52 features). All 48 IDS-SR and BAI items plus 4 demographic variables served as candidate predictors of apathy. Model performance was evaluated with AUC (clinical apathy, SAS >= 14) and R-squared (continuous SAS total). An additional analysis reversed the training direction (NESDA to NESDO) to test generalisability (Supplementary Figure S5).

### APOE analyses

The association between APOE ε4 carrier status and apathy was tested in pooled cross-sectional models (linear and logistic regression) adjusted for age, sex, education, depression severity (IDS-SR), and cohort. Interaction terms (ε4 x age, ε4 x sex, ε4 x depression) were tested to assess potential moderating effects. Longitudinal associations in NESDO were examined using linear mixed models with a time x ε4 interaction term, with and without adjustment for time-varying depression severity (Figure 7). Factor-level analyses tested ε4 associations with each of the three apathy factors (F6, F8, F9) separately. All p-values were FDR-corrected.

## Data availability statement

The data that support the findings of this study are not publicly available due to ethical and legal restrictions on sharing potentially identifying clinical participant information. Data from the Netherlands Study of Depression and Anxiety (NESDA) are available to researchers outside the consortium upon reasonable request and after approval of a formal analysis plan; further information on the data access procedure is available at https://www.nesda.nl/. Data from the Netherlands Study of Depression in Older Persons (NESDO) are similarly available to researchers upon reasonable request and after approval of an analysis plan by the NESDO steering committee; further information is available at https://nesdo.onderzoek.io/. The analyses in this paper were carried out under NESDA data access project DAP2543 and NESDO DAP2676.

## Acknowledgements

S.Z. and M.H. were funded by the Wellcome Trust (226645/Z/22/Z). M.H. was also funded by the National Institute for Health Research (NIHR) Oxford Health Biomedical Research Centre.

We thank the participants of the NESDA and NESDO studies and the cohort teams for their continued work in maintaining these resources. Data access for the present analyses was approved by the NESDA Data Access Committee (reference DAP25-43) and the NESDO Data Access Committee (reference DAP2026-76).

The infrastructure for the NESDA study (www.nesda.nl) is funded through the Geestkracht programme of the Netherlands Organisation for Health Research and Development (ZonMw, grant number 10-000-1002) and is supported by participating universities and mental health care organisations (VU University Medical Center, GGZ inGeest, Leiden University Medical Center, GGZ Rivierduinen, University Medical Center Groningen, Lentis, GGZ Friesland, GGZ Drenthe, Scientific Institute for Quality of Healthcare (IQ healthcare), Netherlands Institute for Health Services Research (NIVEL) and Netherlands Institute of Mental Health and Addiction (Trimbos Institute)).

The Netherlands Study of Depression in Older persons (NESDO) is funded by the Netherlands Organisation for Health Research and Development (ZonMw, grant number 31160004) and is supported by participating universities and mental health care organisations (VU University Medical Center, GGZ inGeest, Leiden University Medical Center, GGZ Rivierduinen, University Medical Center Groningen, Lentis, GGZ Friesland, GGZ Drenthe, Radboud University Nijmegen Medical Centre and Pro Persona).

## Disclosures

All authors declare no financial or non-financial competing interests.

## Notes

### Competing Interest Statement

The authors have declared no competing interest.

### Author Declarations

The study protocols of NESDA and NESDO were approved centrally by the Ethical Review Board of the VU University Medical Center, and subsequently by the local ethical review boards of the participating centres: University Medical Center Groningen and Leiden University Medical Center for NESDA; and Leiden University Medical Center, University Medical Center Groningen, and Radboud University Medical Center Nijmegen for NESDO. Written informed consent was obtained from all participants at the start of baseline assessment.

### Summary of Updates

(1) Title changed to "Apathy precedes worsening depression, but not the reverse". This better reflects the main longitudinal finding. (2) Item-level analysis added, comparing IDS19 and IDS21 against apathy measures to justify the anhedonia operationalisation. (3) Sensitivity analysis added to the cross-lagged panel models, using a 2-item anhedonia composite (IDS19 + IDS21); main conclusions unchanged. (4) Figure 6 (longitudinal results) updated. (5) Supplementary materials updated to reflect the above.

