## Supplementary for "Apathy precedes worsening depression, but not the reverse"

### Depression and anxiety measures are insufficient to capture clinical apathy

From a clinical utility perspective, we investigated whether a dedicated apathy scale is clinically necessary when detailed mood phenotypes are already available. Using minimum redundancy-maximum relevance (mRMR) feature selection followed by elastic net regularisation, we trained prediction models on the older NESDO cohort (N = 388) and validated them on the independent NESDA cohort (N = 1,835). We utilised all 48 IDS-SR and BAI items, alongside four demographic variables, as candidate predictors of apathy.

Feature selection identified four IDS-SR items and one BAI item that best predicted apathy: "reduced general interest" (IDS17), "reduced energy level" (IDS18), "difficulty concentrating" (IDS13), "reduced capacity for pleasure" (IDS19), and "unable to relax" (BAI04). Notably, these are all apathy-adjacent symptoms that straddle the boundary between depression and apathy, rather than core depressive symptoms such as sadness or guilt. At the group level, these five items classified clinical apathy (SAS $\geq$ 14) with acceptable discrimination (AUC = 0.75), and adding the remaining 47 items provided no further improvement (AUC = 0.75; **Supplementary Figure S5A–B**).

Group-level classification, however, masks the poor individual-level prediction. Even the full 52-feature model explained only 27% of continuous apathy variance on cross-cohort validation ($R^2$ = 0.27), while the parsimonious 5-item model explained just 12% ($R^2$ = 0.12; **Supplementary Figure S5C**). This means that 73–88% of individual variation in apathy severity cannot be recovered from depression and anxiety items. A sensitivity analysis reversing the training direction (train NESDA, validate NESDO) yielded somewhat higher performance ($R^2$ = 0.33–0.39; AUC = 0.78–0.82, peaking at 15 features), as expected with a larger training sample, but still left at least 61% of apathy variance unrecovered. In both directions, the majority of clinically relevant apathy variation cannot be reconstructed from standard depression and anxiety items — indicating that apathy must be measured as an independent construct rather than inferred from existing mood instruments.

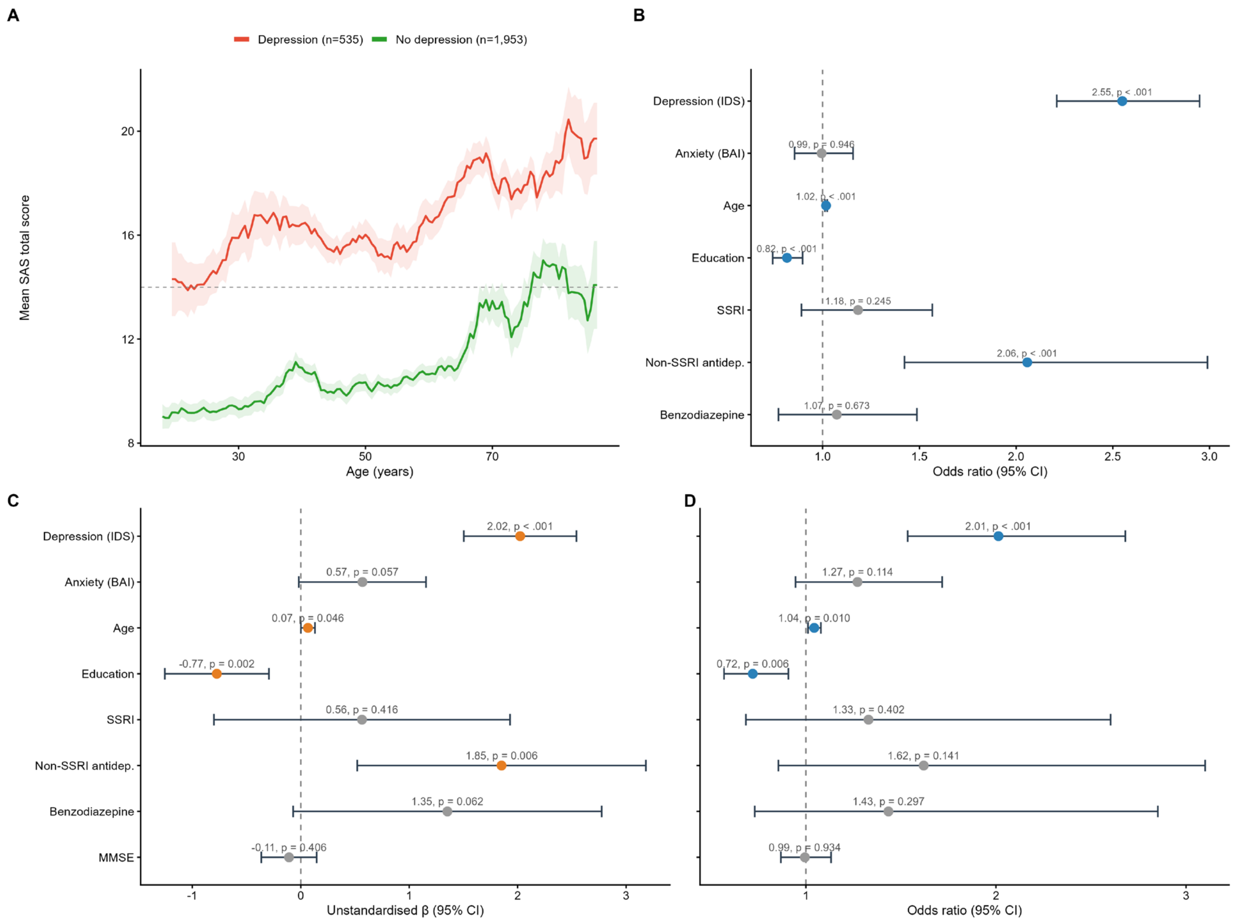


**Supplementary Figure S1: Additional analyses supporting Figure 1.**

(A) Mean apathy score (SAS total) across the adult lifespan by depression status (IDS-SR ≥ 14), using a 5-year sliding window (±2.5 yr; band = ±1 SEM). Dashed line indicates the clinical apathy cutoff (SAS = 14). (B) Odds ratios from logistic regression predicting clinical apathy (SAS ≥ 14; N = 2,469). (C–D) Sensitivity analysis: cognitive function (MMSE) does not confound the age–apathy association. NESDO subsample (N = 465, age 60–93).

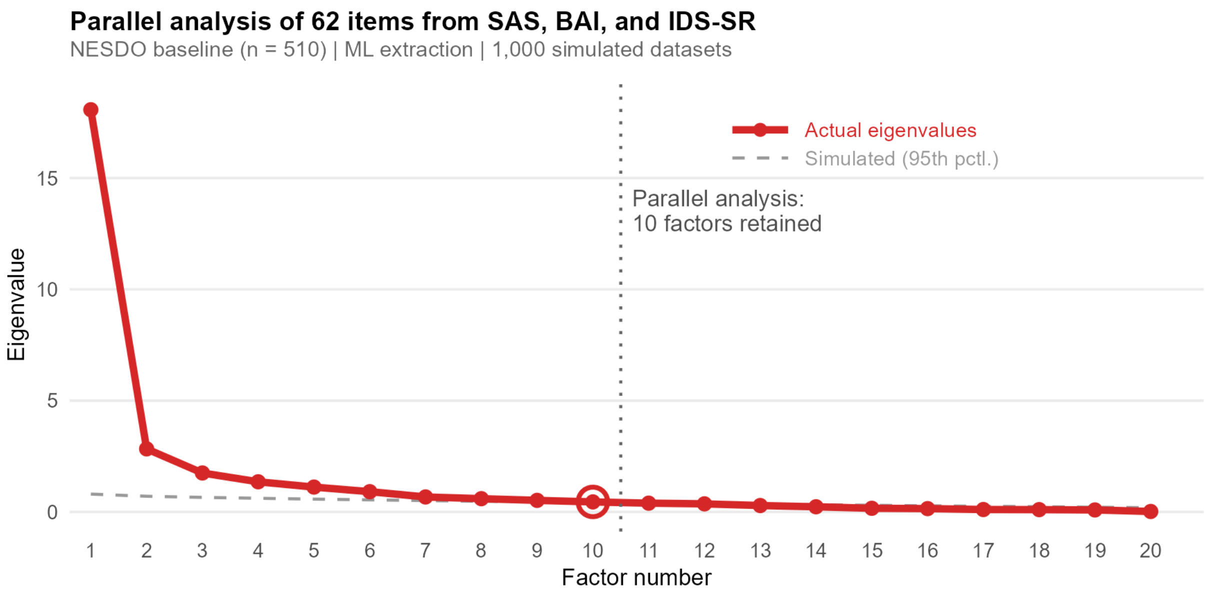


**Supplementary Figure S2: Parallel analysis for determining the number of factors.**

Eigenvalues from the observed correlation matrix (solid line) compared with eigenvalues from 1,000 simulated random datasets (dashed line = 95th percentile). Ten factors met this criterion. NESDO baseline sample (N = 510, 62 items). Maximum likelihood extraction.

### Supplementary Table S1: Complete item-level factor loadings from exploratory factor analysis.

Maximum likelihood extraction with oblimin rotation on 62 symptom items (14 SAS, 21 BAI, 27 IDS-SR) in the NESDO baseline sample (N = 510, age 60–93). Loadings ≥ 0.30 are shown in bold; loadings < 0.30 are suppressed for clarity. h² = communality. See HTML viewer for full interactive table.

| Item | Scale | F1 | F2 | F3 | F4 | F5 | F6 | F7 | F8 | F9 | F10 | h² |
|---|---|---|---|---|---|---|---|---|---|---|---|---|
| AS01 | SAS | | | | | | | | **.86** | | | .77 |
| AS02 | SAS | | | | | | | | **.72** | | | .61 |
| AS03 | SAS | | | | | | | | | | | .29 |
| AS04 | SAS | | | | | | | | | **.53** | | .43 |
| AS05 | SAS | | | | | | | | | **.49** | | .39 |
| AS06 | SAS | | | | | | | | **.35** | **.34** | | .44 |
| AS07 | SAS | | | | | | | | **.35** | **.52** | | .64 |
| AS08 | SAS | | | | **.37** | | | | | | | .57 |
| AS09 | SAS | | | | | | **.36** | | | | | .23 |
| AS10 | SAS | | | | | | **.67** | | | | | .44 |
| AS11 | SAS | | | | | | **.85** | | | | | .67 |
| AS12 | SAS | | | | | | **.53** | | | | | .48 |
| AS13 | SAS | | | | | | **.37** | | | | | .38 |
| AS14 | SAS | | | | | | **.33** | | | | | .43 |
| BAI01 | BAI | | | | | | | | | | | .38 |
| BAI02 | BAI | | | | | **.81** | | | | | | .60 |
| BAI03 | BAI | | | **.47** | | | | | | | | .59 |
| BAI04 | BAI | | **.38** | | | | | | | | | .56 |
| BAI05 | BAI | | **.67** | | | | | | | | | .63 |
| BAI06 | BAI | | | | | | | **.58** | | | | .64 |
| BAI07 | BAI | | | | | | | | | | **.34** | .51 |
| BAI08 | BAI | | | | | | | **.76** | | | | .74 |
| BAI09 | BAI | | **.76** | | | | | | | | | .70 |
| BAI10 | BAI | | **.41** | | | | | | | | | .61 |
| BAI11 | BAI | | **.43** | | | | | | | | | .39 |
| BAI12 | BAI | | | **.90** | | | | | | | | .84 |
| BAI13 | BAI | | | **.86** | | | | | | | | .86 |
| BAI14 | BAI | | **.61** | | | | | | | | | .66 |
| BAI15 | BAI | | **.35** | | | | | | | | | .41 |
| BAI16 | BAI | | **.66** | | | | | | | | | .45 |
| BAI17 | BAI | | **.82** | | | | | | | | | .75 |
| BAI18 | BAI | | | | | | | | | | **.35** | .34 |
| BAI19 | BAI | | | | | | | | | | | .48 |
| BAI20 | BAI | | | | | **.74** | | | | | | .58 |
| BAI21 | BAI | | | | | **.73** | | | | | | .64 |
| IDS01 | IDS | | | | | | | | | | **.30** | .27 |
| IDS02 | IDS | | | | | | | | | | | .09 |
| IDS03 | IDS | | | | | | | | | | **.30** | .24 |
| IDS04 | IDS | | | | | | | | | | **−.43** | .26 |
| IDS05 | IDS | **.64** | | | | | | | | | | .75 |
| IDS06 | IDS | **.55** | | | | | | | | | | .49 |
| IDS07 | IDS | **.47** | **.30** | | | | | | | | | .70 |
| IDS08 | IDS | **.58** | | | | | | | | | | .46 |
| IDS10 | IDS | **.51** | | | | | | | | | | .44 |
| IDS11 | IDS | | | | | | | | | | | .12 |
| IDS12 | IDS | | | | | | | | | | | .09 |
| IDS13 | IDS | **.45** | | | | | | | | | | .49 |

| | | | | | | | | | | | | |
|---|---|---|---|---|---|---|---|---|---|---|---|---|
| IDS14 | IDS | **.45** | | | | | | | | | | .46 |
| IDS15 | IDS | **.50** | | | | | | | | | | .57 |
| IDS16 | IDS | **.42** | | | | | | | | | | .41 |
| IDS17 | IDS | **.50** | | | | | | | | | | .57 |
| IDS18 | IDS | | | | **.57** | | | | | | | .68 |
| IDS19 | IDS | **.51** | | | | | | | | | | .62 |
| IDS20 | IDS | | | | | | | | | | | .32 |
| IDS21 | IDS | **.39** | | | | | | | | | | .39 |
| IDS22 | IDS | **.40** | | | | | | | | | | .38 |
| IDS23 | IDS | | | | **.46** | | | | | | | .55 |
| IDS24 | IDS | | | | | | | | | | **.33** | .51 |
| IDS25 | IDS | **.39** | **.34** | | | | | | | | | .46 |
| IDS26 | IDS | | | | | | | | | | | .23 |
| IDS27 | IDS | **.45** | | | | | | | | | | .32 |
| IDS28 | IDS | | | | **.58** | | | | | | | .67 |

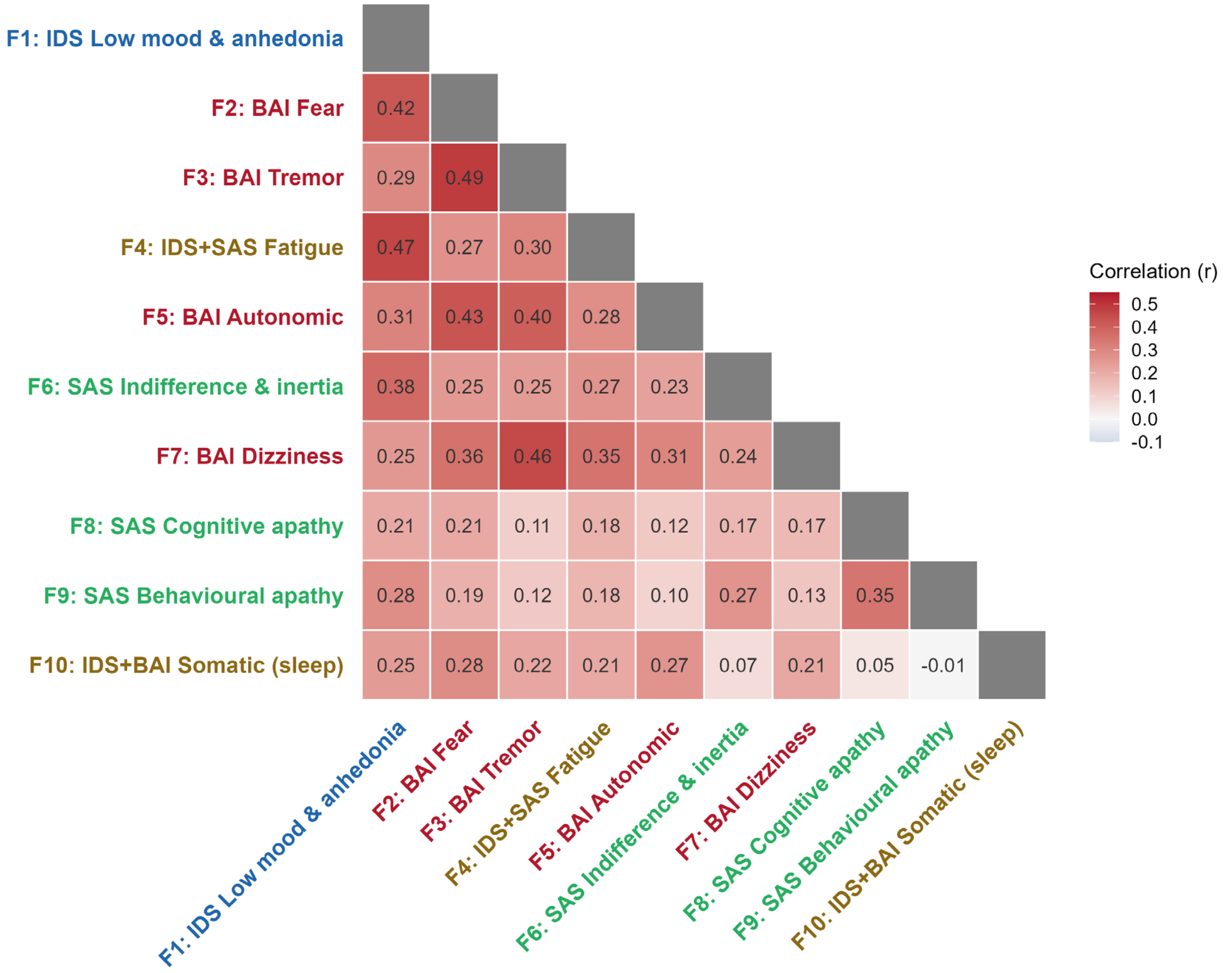


**Supplementary Figure S3: Inter-factor correlation matrix from the 10-factor EFA solution.**

Oblimin-rotated factor correlations (lower triangle) in the NESDO baseline sample (N = 510). Colour intensity reflects correlation magnitude. Axis label colours: green = SAS (apathy), red = BAI (anxiety), blue = IDS (depression), brown = cross-scale.

**Supplementary Table S2: Psychomotor retardation (IDS item 21, "feeling slowed down") correlates primarily with depressive and somatic factors, not apathy.**

Partial Spearman correlations controlling for age, MMSE, and education in the NESDO baseline sample (N = 472).

| Factor | Description | Domain | Partial ρ | p |
|---|---|---|---|---|
| F1 | Depressed mood & anhedonia (excl. IDS21) | Depression | .59 | < .001 |
| F4 | Fatigue | Cross-scale | .44 | < .001 |
| F6 | Indifference & inertia | Apathy | .39 | < .001 |
| F8 | Cognitive apathy | Apathy | .21 | < .001 |
| F9 | Behavioural apathy | Apathy | .26 | < .001 |

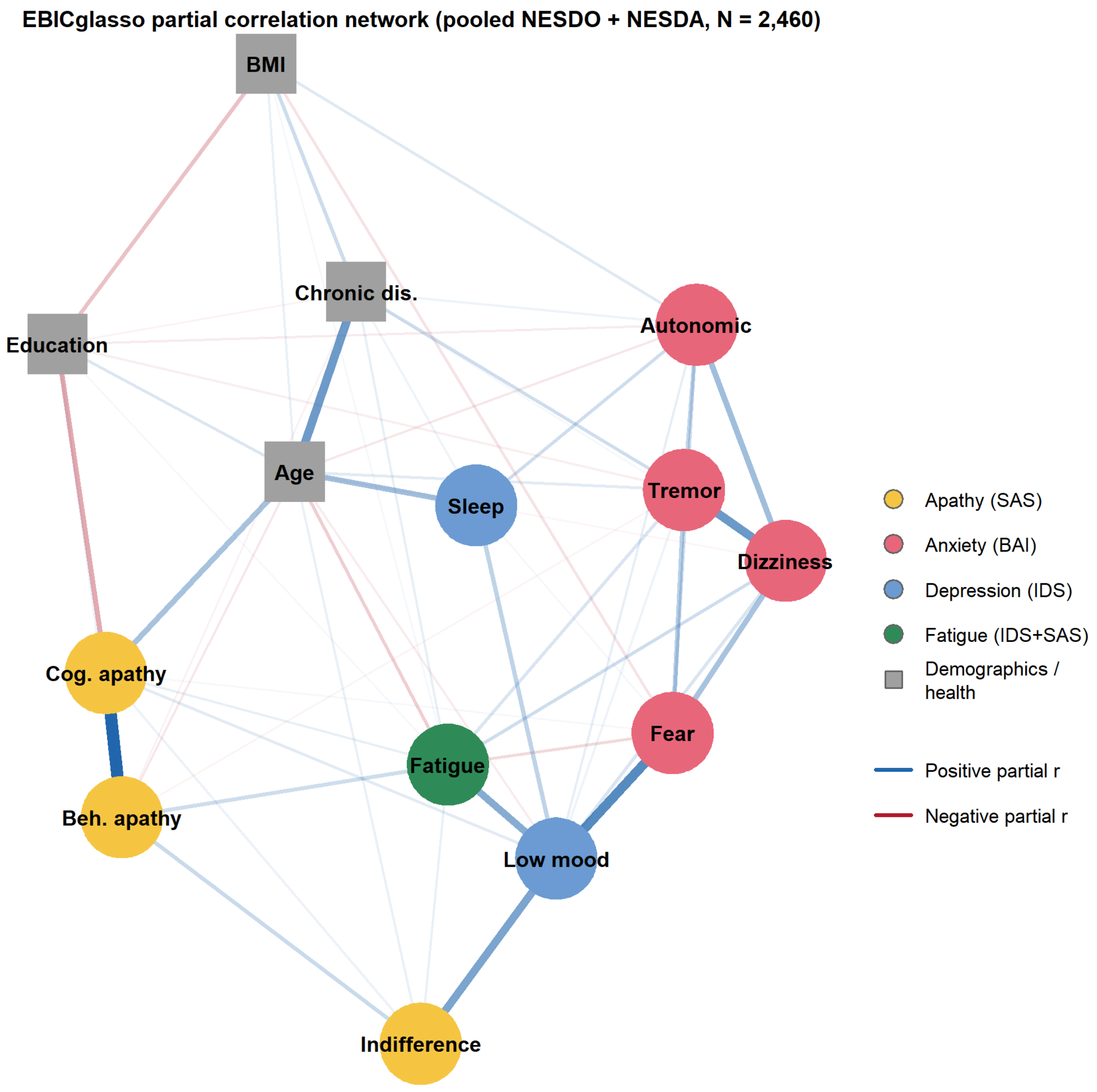


**Supplementary Figure S4: Regularised partial correlation network of symptom factors and clinical variables.**

EBICglasso network estimated on the pooled NESDO + NESDA sample (N = 2,460; 14 nodes; 69 non-zero edges of 91 possible). The primary cross-domain bridge connects depressed mood (F1) to indifference and inertia (F6; partial r = 0.328). Within the apathy cluster, the cognitive–behavioural apathy edge (F8–F9; partial r = 0.551) is the strongest in the entire network.

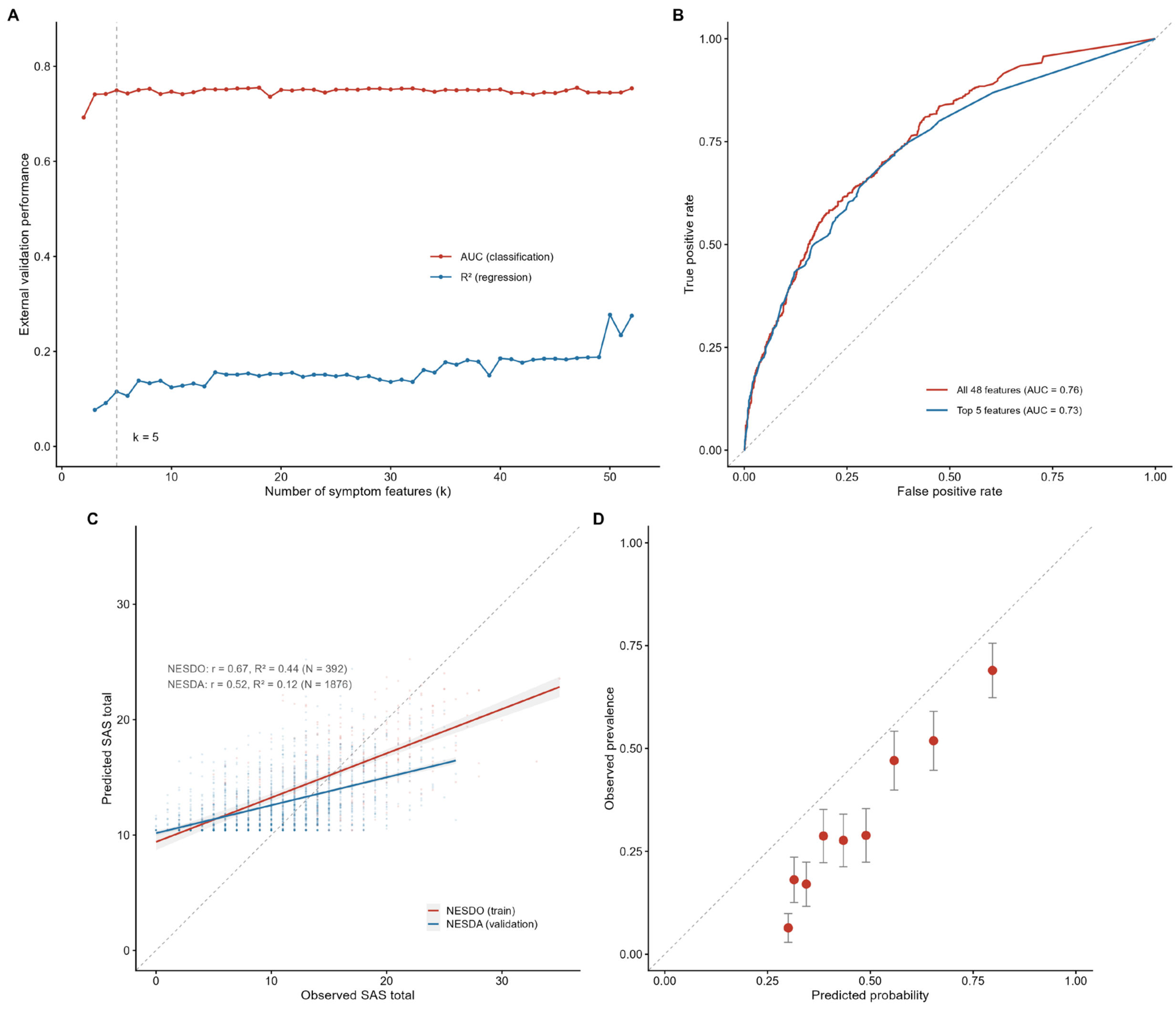


**Supplementary Figure S5: Can clinical apathy be inferred from depression and anxiety measures alone?**

Minimum redundancy–maximum relevance (mRMR) feature selection followed by elastic net regularisation, trained on NESDO (N = 388) and validated on NESDA (N = 1,835). (A) External validation performance as a function of mRMR-selected features. (B) ROC curves. (C) Predicted vs observed SAS total. (D) Calibration plot.

**Supplementary Table S3: Cross-lagged panel model results: bidirectional effects between apathy and symptom dimensions.**

Standardised cross-lagged coefficients (β) from bivariate CLPMs in the NESDO cohort (N = 374). Bonferroni-corrected threshold for 12 tests: α = 0.004.

| Model | Direction | Period | β | SE | p | Sig |
|---|---|---|---|---|---|---|
| Anhedonia | Symptom → Apathy | Baseline → 2yr | .009 | .056 | .868 | |
| Anhedonia | Symptom → Apathy | 2yr → 6yr | .132 | .050 | .008 | ** |
| Anhedonia | Apathy → Symptom | Baseline → 2yr | .257 | .063 | < .001 | *** |
| Anhedonia | Apathy → Symptom | 2yr → 6yr | .199 | .060 | < .001 | *** |
| Depressed mood | Symptom → Apathy | Baseline → 2yr | .109 | .057 | .055 | |
| Depressed mood | Symptom → Apathy | 2yr → 6yr | .063 | .050 | .214 | |
| Depressed mood | Apathy → Symptom | Baseline → 2yr | .105 | .062 | .088 | |
| Depressed mood | Apathy → Symptom | 2yr → 6yr | .180 | .057 | .001 | ** |
| Anxiety | Symptom → Apathy | Baseline → 2yr | .052 | .057 | .359 | |
| Anxiety | Symptom → Apathy | 2yr → 6yr | .039 | .048 | .412 | |
| Anxiety | Apathy → Symptom | Baseline → 2yr | .063 | .058 | .276 | |
| Anxiety | Apathy → Symptom | 2yr → 6yr | .089 | .046 | .056 | |

### Supplementary Table S4: Latent growth curve model: baseline symptom predictors of apathy trajectory.

Standardised regression coefficients (β) from a LGCM predicting the intercept (baseline level) and slope (rate of change) of apathy (SAS total) over 6 years in NESDO (N = 374). Baseline mood symptoms predicted concurrent apathy level but not its rate of change.

| Growth factor | Predictor | β | SE | p | Sig |
|---|---|---|---|---|---|
| Intercept | Anxiety (BAI F2) | .203 | .059 | < .001 | *** |
| Intercept | Anhedonia (IDS19) | .134 | .060 | .025 | * |
| Intercept | Depressed mood (IDS composite) | .283 | .061 | < .001 | *** |
| Intercept | MMSE | −.016 | .046 | .734 | |
| Slope | Anxiety (BAI F2) | −.012 | .009 | .200 | |
| Slope | Anhedonia (IDS19) | −.004 | .010 | .646 | |
| Slope | Depressed mood (IDS composite) | −.007 | .010 | .497 | |
| Slope | MMSE | −.002 | .007 | .796 | |

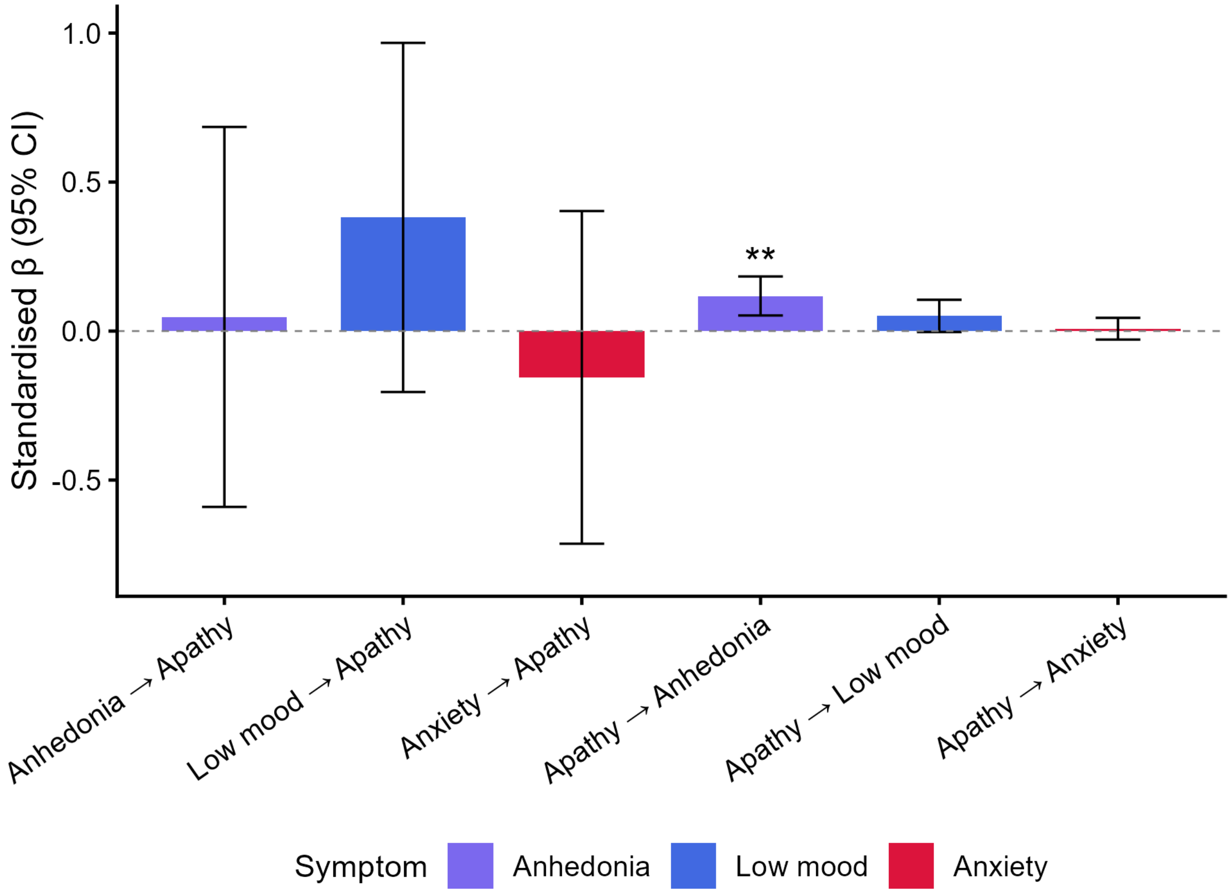


**Supplementary Figure S6: Reverse prediction simultaneous 4-variable cross-lagged panel model.**

Standardised cross-lagged coefficients (β with 95% CI) from a simultaneous CLPM modelling apathy, anhedonia, depressed mood, and anxiety together in NESDO (N = 374; 2-year → 6-year period). When all symptom dimensions compete simultaneously, only the apathy → anhedonia path reaches significance. No symptom dimension significantly predicted later apathy.

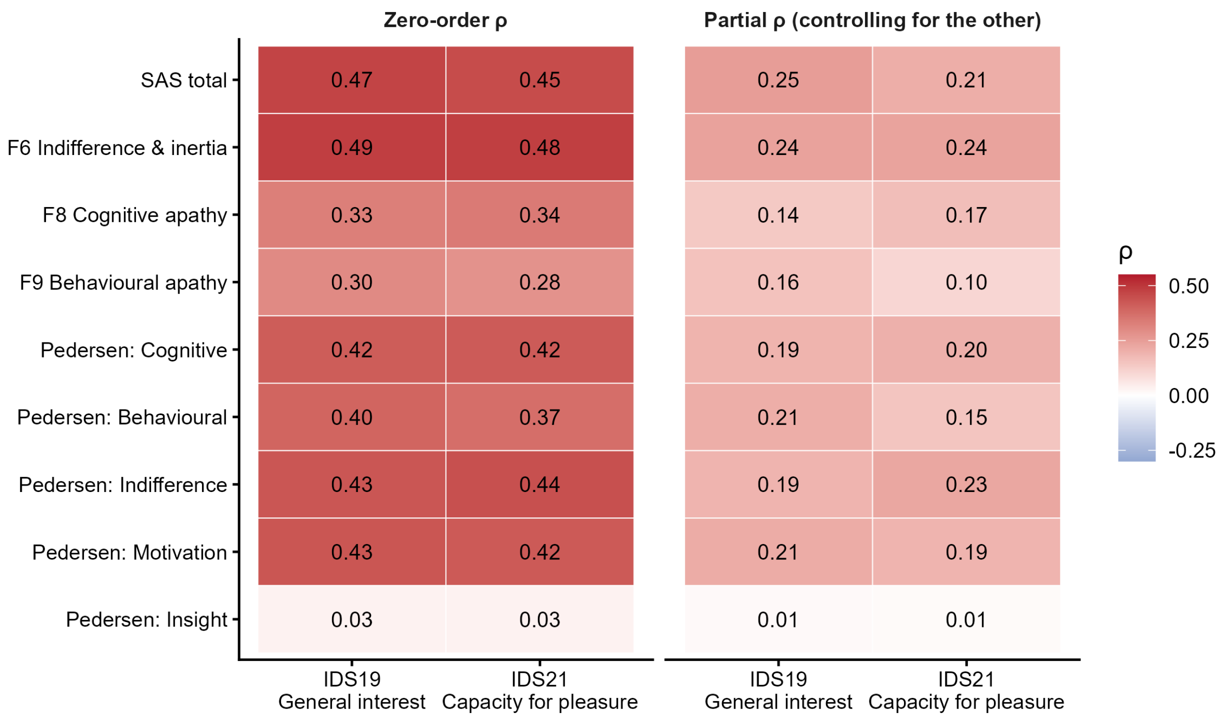


**Supplementary Figure S7: IDS-SR item 19 (general interest) versus item 21 (capacity for pleasure) coupling with apathy domains.**

Pooled NESDO baseline + NESDA Wave E sample (N = 2,609). Left panel: zero-order Spearman ρ. Right panel: partial Spearman ρ controlling for the other item. IDS19 was significantly more strongly correlated with apathy than IDS21 was for SAS total (Steiger Z = +3.13, FDR = .005), behavioural apathy (F9: Z = +3.67, FDR = .001) and Pedersen Behavioural (Z = +4.22, FDR < .001), IDS21 was more strongly correlated than IDS19 only with Pedersen Indifference (Z = −2.54, FDR = .025), the apathy domain closest to consummatory anhedonia. The two items therefore capture different aspects of motivation and pleasure, supporting IDS21 as the cleaner single-item anhedonia indicator in the main analyses.

**Supplementary Table S5: Bivariate cross-lagged panel models using a 2-item anhedonia composite (IDS items 19 + 21).**

Standardised cross-lagged coefficients (β) from bivariate CLPMs in the NESDO cohort (N = 292), using a 2-item anhedonia composite (mean of IDS-SR item 19 "general interest" and item 21 "capacity for pleasure"). Compared against the main single-item analysis (Supplementary Table S3), all four anhedonia paths are within ~0.01 of the main-text estimates, so the asymmetric apathy → anhedonia precedence does not depend on the single-item indicator. Bonferroni-corrected threshold for 12 tests: α = 0.004.

| Model | Direction | Period | β | SE | p | Sig |
|---|---|---|---|---|---|---|
| Anhedonia (2-item) | Symptom→Apathy | Baseline → 2yr | .022 | .059 | .710 | |
| Anhedonia (2-item) | Apathy→Symptom | Baseline → 2yr | .249 | .066 | < .001 | *** |
| Anhedonia (2-item) | Symptom→Apathy | 2yr → 6yr | .129 | .052 | .012 | * |
| Anhedonia (2-item) | Apathy→Symptom | 2yr → 6yr | .197 | .057 | < .001 | *** |
| Low mood | Symptom→Apathy | Baseline → 2yr | .109 | .057 | .055 | |
| Low mood | Apathy→Symptom | Baseline → 2yr | .105 | .062 | .088 | |
| Low mood | Symptom→Apathy | 2yr → 6yr | .063 | .050 | .214 | |
| Low mood | Apathy→Symptom | 2yr → 6yr | .180 | .057 | .001 | ** |
| Anxiety | Symptom→Apathy | Baseline → 2yr | .052 | .057 | .359 | |
| Anxiety | Apathy→Symptom | Baseline → 2yr | .063 | .058 | .276 | |
| Anxiety | Symptom→Apathy | 2yr → 6yr | .039 | .048 | .412 | |
| Anxiety | Apathy→Symptom | 2yr → 6yr | .089 | .046 | .056 | |

**Supplementary Table S6: Full 4-variable cross-lagged panel model using a 2-item anhedonia composite (IDS items 19 + 21).**

All 24 cross-lagged paths from the simultaneous 4-variable CLPM (apathy, anhedonia, depressed mood, anxiety) in the NESDO cohort (N = 292), with FDR-adjusted p-values. The apathy → anhedonia path remains the strongest cross-lagged path in the 2-to-6-year period (β = 0.187, FDR = .014); the reverse anhedonia → apathy path does not survive FDR correction (β = 0.138, FDR = .12).

| Period | From | To | β (std) | p | FDR | Sig |
|---|---|---|---|---|---|---|
| T1→T2 | Anhedonia | Apathy | −.045 | .507 | .727 | |
| T1→T2 | Sadness | Apathy | .117 | .086 | .207 | |
| T1→T2 | Anxiety | Apathy | .020 | .755 | .916 | |
| T1→T2 | Apathy | Anhedonia | .171 | .012 | .059 | |
| T1→T2 | Sadness | Anhedonia | .275 | < .001 | .007 | ** |
| T1→T2 | Anxiety | Anhedonia | .032 | .640 | .853 | |
| T1→T2 | Apathy | Sadness | .086 | .191 | .353 | |
| T1→T2 | Anhedonia | Sadness | −.082 | .257 | .440 | |
| T1→T2 | Anxiety | Sadness | .184 | .006 | .037 | * |
| T1→T2 | Apathy | Anxiety | −.009 | .891 | .916 | |
| T1→T2 | Anhedonia | Anxiety | −.016 | .810 | .916 | |
| T1→T2 | Sadness | Anxiety | .217 | .002 | .014 | * |
| T2→T3 | Anhedonia | Apathy | .138 | .034 | .116 | |
| T2→T3 | Sadness | Apathy | −.019 | .774 | .916 | |
| T2→T3 | Anxiety | Apathy | .006 | .916 | .916 | |
| T2→T3 | Apathy | Anhedonia | .187 | .002 | .014 | * |
| T2→T3 | Sadness | Anhedonia | .010 | .898 | .916 | |
| T2→T3 | Anxiety | Anhedonia | .040 | .515 | .727 | |
| T2→T3 | Apathy | Sadness | .119 | .049 | .145 | |
| T2→T3 | Anhedonia | Sadness | .121 | .099 | .215 | |
| T2→T3 | Anxiety | Sadness | .121 | .054 | .145 | |
| T2→T3 | Apathy | Anxiety | .072 | .169 | .338 | |
| T2→T3 | Anhedonia | Anxiety | −.064 | .312 | .499 | |
| T2→T3 | Sadness | Anxiety | .150 | .022 | .089 | |
